# A Cluster-Randomized Trial to Compare Effectiveness and Cost-effectiveness of Cash Plus Interventions in Preventing Child Wasting in Somalia: An Evidence-Based Methodology

**DOI:** 10.1101/2025.02.21.25322679

**Authors:** Shelley Walton, Kemish Kenneth Alier, Sydney Garretson, Samantha Grounds, Qundeel Khattak, Marina Tripaldi, Fabrizio Loddo, Said Aden Mohamoud, Mohamud Ali Nur, Sadiq Abdiqadir, Emily Mitchell, Mohamed Billow Mahat, Maimun Gure, Dahir I. Jibril, Dahir Gedi, Michael Ocircan, Meftuh Omer, Abdullahi Arays, Abdifitah Ahmed Mohamed, Adam Abdulkadir Mohamed, Nadia Akseer

## Abstract

**Background:** Somalia is a conflict, flood, and drought prone country with high rates of food insecurity and child wasting. Save the Children partnered with Johns Hopkins University to study the most effective and cost-effective combinations of assistance to prevent acute malnutrition among PLW and CU5 in a six-month humanitarian program.

**Objectives:** This study implemented an cluster-randomized controlled trial using adaptive design methodology to: (A) Estimate and compare wasting incidence and prevalence of CU5 and their mothers receiving on a monthly basis either cash (Arm 1), cash + social and behavior change communication (Arm 2), or cash+ top-up cash (Arm 3); after 3 months and 6 months; (B) Calculate the costs and cost-effectiveness of the different intervention arms; (C) Understand perspectives and experiences of mothers and fathers of CU5 beneficiaries; (D) Monitor the functionality of markets and availability and prices of foods. This paper presents the approach that was designed and implemented to study these objectives.

**Methodology:** This study employed a mixed-methods approach with a quantitative component conducted at three timepoints to collect anthropometric measurements and household survey data. Primary outcomes, such as child and maternal wasting, were assessed using standardized WHO criteria. Additional data on food security, maternal and child health, and household conditions were collected to evaluate immediate, underlying, and basic causes of malnutrition. Cost analyses evaluated programmatic and societal costs of the intervention. An adaptive trial design was implemented, allowing the methodology to evolve as new challenges emerged.

**Conclusion:** This trial applied an adaptive mixed methods design to evaluate the effectiveness and cost-effectiveness of cash assistance interventions in Somalia, overcoming complex humanitarian operational challenges. Strong partnerships and flexible trial design allowed us to adjust to unpredictable events and maintain research rigor. These findings highlight the value of adaptive designs and mixed methods for improving child nutrition outcomes in complex settings.

**Registration:** The cluster-RCT is registered at ClinicalTrials.gov, ID: NCT06642012.

## 1. Background

In Somalia, acute malnutrition case admissions among CU5 have risen dramatically in recent years.[1] Due to climate shocks (prolonged drought and flooding), persistent conflict, and global supply and price shocks, millions of people in Somalia are suffering from food insecurity and experiencing one of the most complex humanitarian crises.[2] The Food Security and Nutrition Analysis Unit’s (FSNAU’s) recent survey showed that at least 1.5 million children under five (42.8% of the total under-five children in Somalia) and 4.3 million people overall are projected to be acutely malnourished from August 2023 through July 2024, with a projected 330,630 children being severely malnourished.[3]

Countries like Somalia rely heavily on humanitarian initiatives, such as food and cash transfers, specialized nutritious foods, and nutrition education to combat malnutrition. However, there is limited evidence on the most effective, cost-effective, and acceptable approaches in humanitarian crisis settings. Evidence gaps identified by Elrha evidence reviews in 2015[4] and 2022[5], a research agenda by the World Health Organization (WHO) in 2018[6], and research questions emphasized by the United Nations International Children’s Emergency Fund (UNICEF) in 2021[7] highlight the need for further research to explore 1) cash and voucher assistance (CVA) duration and amount, 2) cost-effectiveness design, and 3) evidence generation in humanitarian settings. These gaps underscore the critical need for targeted research to inform policy and practice, ensuring that humanitarian interventions are both effective and cost-effective in addressing malnutrition in crisis-affected populations.

The unpredictable context of humanitarian settings due to climate or conflict-related events can make it challenging for researchers to study these questions. There is an evidence gap in study methodologies and approaches, especially for mixed-methods impact evaluations and cost analyses, that can provide reliable data to answer these questions under the variable conditions often present in crisis settings.[8–12] Adaptive design methods in humanitarian settings provide a flexible approach where interventions or programs are modified based on real-time data and feedback gathered from the study setting and community throughout the project.[13–15] Adaptive designs allow research and implementers to better respond to changing needs and circumstances in dynamic humanitarian crises. Methodologies that adapt to complex research environments, highlight lessons learned and best practices for researchers, and fill evidence gaps are critical to publish to support future research in humanitarian settings.

This trial was a prospective, cluster-randomized controlled trial following CU5 and PLW to assess the effectiveness and cost-effectiveness of combinations and durations of monthly cash assistance in preventing severe acute malnutrition (SAM) and moderate acute malnutrition (MAM) in a 6-month humanitarian program. The trial was conducted in the Bay and Hiran regions of Somalia, which experience some of the highest rates of wasting in the country.[1,16] This three-arm trial with randomization at the village level used a mixed methods approach, including both quantitative and qualitative data collection and analyses and cost-effectiveness analyses.

Specifically, the primary aims of this trial were to:

A. To estimate and compare wasting incidence and prevalence of children 6-59 months-old and their mothers receiving on a monthly basis either cash (Arm 1 – control), cash + social and behavior change communication (SBCC) (Arm 2), or cash+ top-up cash (Arm 3); after 3 months and 6 months of cash assistance
B. To calculate the costs and cost-effectiveness of the different intervention arms
C. To understand perspectives and experiences of mothers and fathers of CU5 beneficiaries of the cash program
D. To monitor the functionality of markets and availability and prices of foods in the study locations

This study presents and discusses the adaptive evidence-based methodology used in this trial to study combinations of cash interventions in a humanitarian setting.

The methodology was designed to address the evidence gap and inform global guidance on the duration of cash for nutrition programs, the amount of cash to prevent acute malnutrition, and the optimal combinations of interventions in cash for nutrition programs. The published research methodology, study instruments, and lessons learned will support future research in highly variable humanitarian settings and inform future cash for nutrition programs in Somalia and beyond.

## 2. Approach

### 2.1. Trial Setting

The two trial sites were Baidoa and Beledweyne, the capitals of the Bay and Hiran regions, respectively, and were primarily rural.[17,18] Bay and Hiran were selected as trial sites not only due to existing SC BHA CashPlus for Nutrition programming, but also due to their high global acute malnutrition (GAM) and severe acute malnutrition (SAM) rates, with GAM rates of 18.3% and SAM rates of 3.6% in Beledweyne district, and GAM rates of 19.1% and SAM rates of 4.9% in Baidoa-Buurhakaba livelihood zones.[3] Bay and Hiran are located in the southwest region of Somalia. In 2021, the population of Bay was around 1.1 million people, while Hiran had an estimated 427,124 people.[19,20]

Both Bay and Hiran have been greatly affected by climate crises (droughts, floods), instability, and conflict that have affected livelihoods and contributed to the displacement of thousands.[21] Over the years, such insecurities have contributed to the displacement of over 25,000 people from Bay and over 250,000 people from Hiran from July 2021 to November 2022. [22–25] In 2023, Hiran received an influx of nearly half a million new IDPs, and Bay received 219,000.[25] Roughly 25% of the country’s IDP sites are around the Baidoa area in Bay, and these sites experience some of the highest levels of food insecurity in Somalia.[26] In the first quarter of 2023, roughly 750,000 individuals in Bay suffered from IPC level 3 (crisis) or level 4 (emergency) food insecurity.[1]

### 2.2. Conceptual Framework

The UNICEF Conceptual Framework on Maternal and Child Nutrition categorizes the determinants of malnutrition into three groups: immediate, underlying, and basic/enabling causes.[27,28] This evidence-based conceptual framework informed the trial design and both quantitative and qualitative data collection and analysis (Figure S1).

### 2.3. Study Design

This adaptive trial used a mixed methods design inclusive of a difference-in-difference approach for advanced quantitative analysis, qualitative data collection (via focus group discussions), market monitoring surveys and detailed cost, cost efficiency and cost-effectiveness analysis. The research team utilized an explanatory sequential design to integrate the qualitative study components, collecting and analyzing quantitative data first, followed by qualitative data. We then used the qualitative data to help explain and contextualize the quantitative findings.

### 2.4. Trial Methodology Development and Adaptive Design

This trial methodology was co-developed by researchers at Johns Hopkins University, program staff at Save the Children International and Save the Children Somalia, and other global experts on nutrition assistance in humanitarian settings through iterative consultations. The methods described in this paper are the final version of the trial design.

An adaptive design for the trial was adopted to account for the variable humanitarian setting and to ensure the validity and integrity of data collection.[29] For example, severe flooding occurred throughout Somalia and in Bay and Hiran during the trial, affecting almost 2.5 million people and displacing over one million individuals.[24] The flooding resulted in displacement and disrupted access to research sites, affecting data collection. However, the trial team was able to adapt and respond to the crisis in a way that maintained the integrity of the research. The timeline of conflict and climate-related shocks to the study setting is found in Figure 1 below.

**Figure 1.**
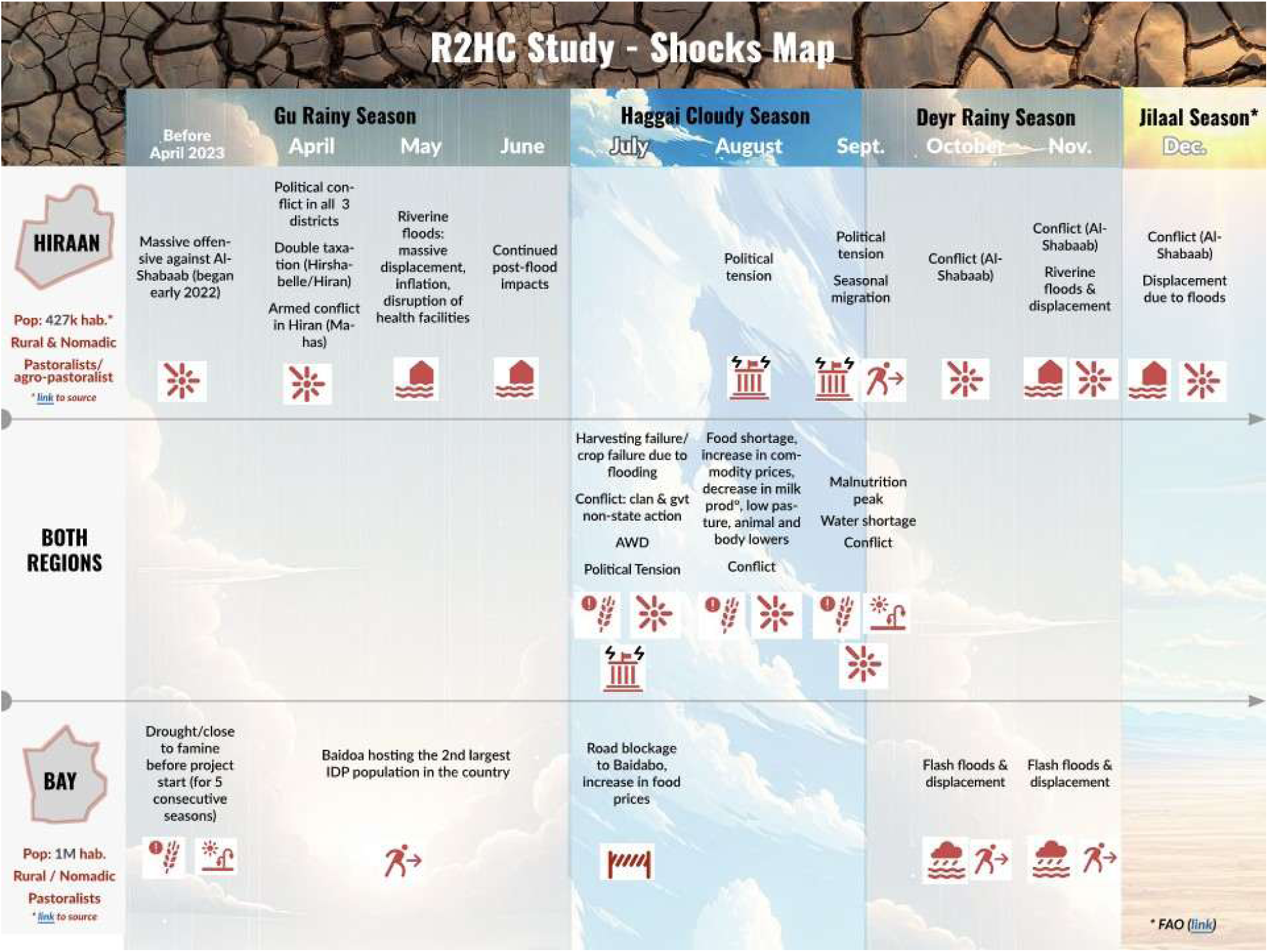
Bay and Hiran 2023 Shocks Map.

### 2.5. Trial Design

The prospective cluster-RCT was designed to study combinations of cash assistance across three study arms. The base cash transfer amounts were established by the National Cash Working Group based on the Minimum Expenditure Basket (MEB). Cash amounts were calculated to meet 80% of a person’s energy needs (kcals) per month and were based on the average number of household members; six to seven members in the Somalia context.[30] Amounts distributed considered fluctuations in food prices due to conflict, climate shocks, and inflation. They allowed for basic food purchases like staple grains, legumes, oil, and vegetables, and occasional purchases of fresh fruits, dairy products, and meat products. The unconditional cash (CVA) for food assistance was designed to increase purchasing power, thereby meeting the immediate needs of most food-insecure people in the target regions. This integrated approach also aimed to prevent child malnutrition and wasting relapse.

The combinations of assistance studied in three separate arms were:

- **Arm 1:** UCT to targeted families – 1 mobile transfer per month for 6 consecutive months. Households in Bay received $90 per month and households in Hiran received $70 per month.
- **Arm 2:** UCT with SBCC – SBCC package included interpersonal communication, such as one-to-one consultations for mothers of CU5, bi-monthly group sessions of approximately 45-60 minutes through the Mother-to-Mother Support Groups on key topics related to nutrition, health, and community awareness raising campaigns (additional details in Annex S2). Households in Bay received $90 per month and households in Hiran received $70 per month.
- **Arm 3:** UCT with additional monthly nutrition-cash top up – 1 per month for 6 months. Households in Bay received $90 plus an additional $35 top-up ($125 total) per month and households in Hiran received $70 plus an additional $35 top-up ($105 total) per month.

The program was implemented from June to November 2023. Baseline quantitative data collection occurred in May 2023, midline occurred in September 2023, and endline occurred in December 2023. Qualitative data were collected in January 2024.

### 2.6. Trial Population and Sampling Methods

The trial participants comprised mothers, fathers and their children (CU5) whose households were enrolled in the BHA CashPlus for Nutrition program.

For CU5 and their mothers, the study inclusion and exclusion criteria are found in Table 2 below.

**Table 1.** Trial Inclusion and Exclusion Criteria.

| Study Participant | Inclusion Criteria | Exclusion Criteria |
| --- | --- | --- |
| Mothers of children 6 to 59 months | • Enrolled in Save the Children CashPlus for Nutrition program<br>• Have a child that is 6-59 months of age at baseline<br>• Subject who is voluntarily willing to participate by signing the consent form | • Currently receiving treatment for MAM* or SAM**<br>• Had an episode of severe wasting (SAM) in the past 12 months |
| Children 6-59 months | • Enrolled in Save the Children CashPlus for Nutrition program<br>• 6-59 months of age at baseline<br>• Their mother is enrolled in the study | • Receiving treatment for MAM or SAM at baseline<br>• Had an episode of severe wasting (SAM) in the past 12 months |

\*Moderate acute malnutrition. \*\*Severe acute malnutrition.

**Table 2.** Primary Outcome Definitions for CU5 and Mothers.

| ***Malnutrition Guidelines for CU5*** |  |
| --- | --- |
| Without Acute Malnutrition | MUAC<sup>1</sup> ≥ 125 mm <u>and</u> WHZ<sup>2</sup> ≥ -2 SD, without edema |
| With Moderate Acute Malnutrition (MAM) | 115 mm ≤ MUAC $<$ 124.9 mm and/or -3 SD ≤ WHZ $<$ -2 SD, without edema |
| With Severe Acute Malnutrition (SAM) | MUAC $<$ 115mm, and/or WHZ $<$ -3 SD, and/or presence of edema |
| ***Stunting Guidelines for CU5*** |  |
| Without Stunting | HAZ<sup>3</sup> $>$ -2 SD |
| Moderate Stunting | -3 SD $<$ HAZ ≤ -2SD |
| Severe Stunting | HAZ ≤ -3 SD |
| ***Malnutrition Guidelines for Mothers of CU5*** |  |
| Overweight | MUAC $>$ 300mm |
| Normal | MUAC ≥ 230 mm |
| With Moderate Acute Malnutrition (MAM) | MUAC $<$ 230 mm |

<sup>1</sup>MUAC: Mid-upper arm circumference
<sup>2</sup>WHZ: Weight-for-height z-score
<sup>3</sup>HAZ: Height-for-age z-score

Prior to baseline data collection, the SC Somalia project team provided a household beneficiary register for the CashPlus for Nutrition program to the research team. The research team reviewed the registration details to generate a list of all CU5 mothers with their CU5s that met the inclusion and exclusion criteria. This produced the sampling frame from which the study sample was selected.

Initially 44 total clusters (villages) were assessed for eligibility. 11 clusters were excluded because these clusters were located in heavily flood-prone areas where households easily migrate, which would have impacted the overall sample size and data collection. The sample size estimation was based on the primary outcome for the CU5, feasibility/logistics in study settings, and a 7% minimum detectable difference in post-intervention prevalence of wasting using: 1) 20% baseline wasting (figures are transient and based on the higher prevalence of the two regions); 2) intra-class correlation coefficient (ICC) from earlier studies (0.02)[31,32]; average cluster size (150 HHs); 4) number of clusters (33 total, 11 per arm); 5) 5% significance and 80% power; and 6) CU5 per household (1.3 children). The initial sample required was 410 HHs per intervention group or 533 children. Accounting for 15% attrition, the final required sample size was 471 HHs or 613 children per arm. 33 clusters were randomized to the three study arms. Each cluster ranged in size from 55 to 431 households, with an average cluster size of 140-158 households. 4,838 total households were evaluated for eligibility against the trial’s inclusion and exclusion criteria. 3,348 households were excluded because they did not have CU5 in the household or did not meet one or more exclusion criteria. 1,490 total households were enrolled in the study. The final child sample was 1,894 children.

Household and mothers of CU5 were selected from health facility registries and were then contacted by phone for study recruitment and mobilization for data collection. Figure 2 shows the trial recruitment and data collection sample for each study timepoint. Table S3 also shows a table of subject recruitment for all study activities.

**Figure 2.**
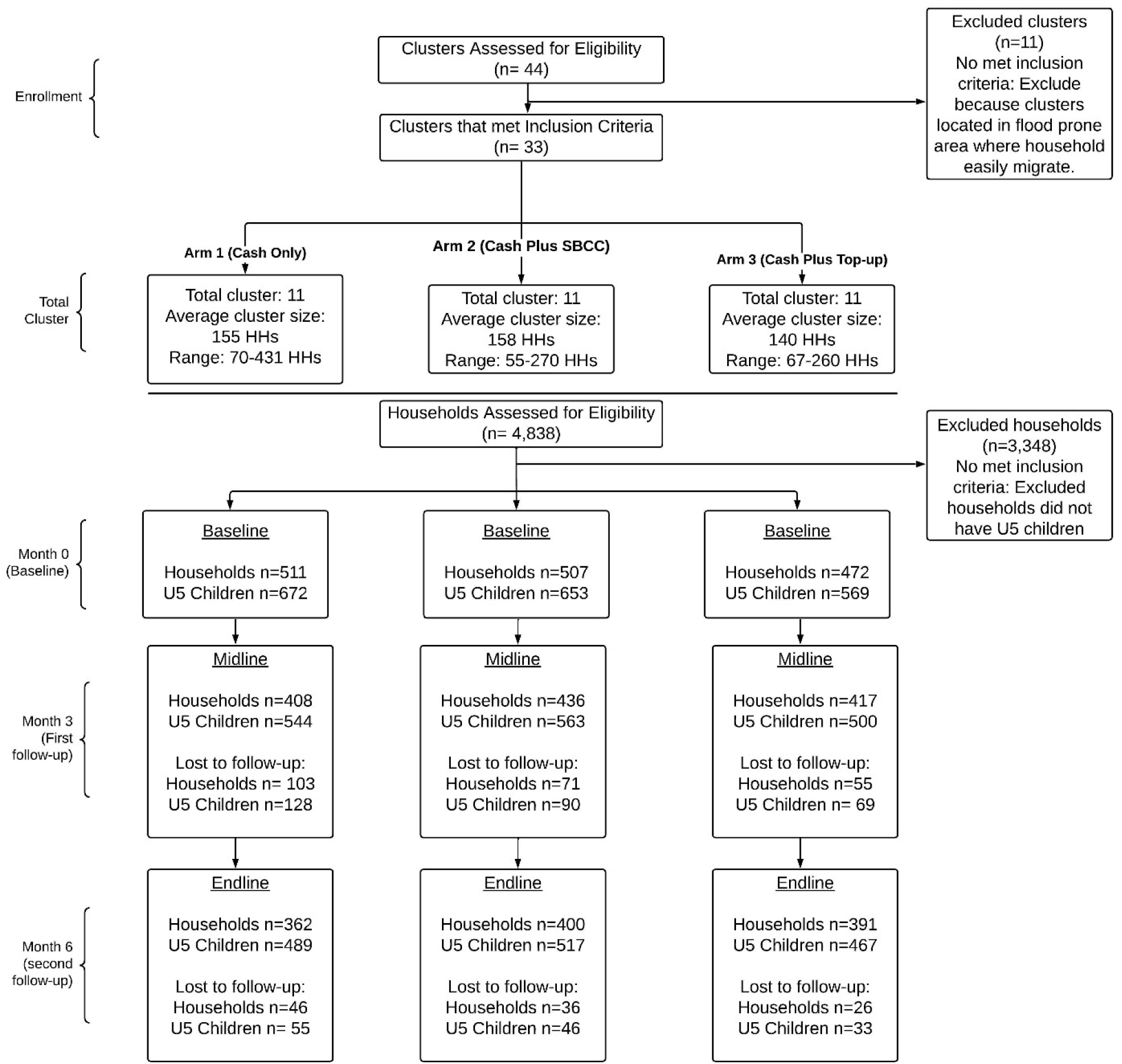
Trial Recruitment Consort Diagram.

## 3. Quantitative Approach

Once enrollment criteria were confirmed and informed consent was obtained, a team of trained enumerators (in pairs) conducted the household survey and collected anthropometric data for the CU5 mother and the youngest CU5 in the household. Anthropometric measures and household surveys were administered to 1490 households at baseline, 1261 households at midline, and 1153 households at endline.

### 3.1. Outcomes

The primary outcomes of interest were child and maternal wasting. A secondary outcome of interest was child stunting. Standard methods for anthropometric measurements were used based on the SMART methodology.[33] At each study timepoint, the CU5 were classified as:

If a CU5 or CU5 mother was identified as wasted during baseline, midline or endline, he/she was referred to the nearest health center that offered wasting treatment; however, he/she remained in the study cohort for the duration of the study.

The primary child nutrition outcomes were assessed using MUAC, WHZ, and presence of edema to generate categorical indicator variables that align with the 2013 WHO Child Growth Standards criteria.[34] These indicator variables were used to compare child malnutrition at each timepoint to measure changes in wasting over the life of the study and between study arms to examine benefits of each intervention type. Intergenerational nutrition outcomes were assessed using size at birth of the youngest child (collected at baseline). Maternal nutrition outcomes were assessed using MUAC to generate categorical indicator variables (malnourished, normal, and overweight), and the proportion of malnourished mothers were compared between study arms and at each data collection timepoint.[35]

### 3.2. Exposures

Mothers completed household quantitative surveys at baseline, midline (3 months post-baseline), and endline (6 months post-baseline) to establish benchmarks and explore changes related to the intended outcomes and determinants. These questionnaires were translated to Somali and administered to all study participants (mothers of CU5). There are slight variations in each questionnaire, such as incorporating questions around displacement due to the flooding that occurred in the endline questionnaire, and all questionnaire versions are included in the supplementary materials (Tables S4-S6).

The following data were collected during the household survey at each time point and were guided by the UNICEF conceptual framework of maternal and child nutrition:[27,28]

Immediate Causes: Child diet was measured using global infant and young child feeding (IYCF) indicators such as exclusive breastfeeding, minimum dietary diversity (MDD), minimum meal frequency (MMF), minimum acceptable diet (MAD), and consumption of different food groups (fruits/vegetables, protein sources, etc.). Child health history was tracked over the life of the study; mothers were asked about the CU5’s episodes of diarrhea, pneumonia, fever or other illnesses in the last two weeks. Maternal age, MUAC, and pregnancy status were also considered immediate causes.

Underlying Causes: Household food insecurity was also evaluated using the Household Hunger Scale (HHS); the Food Consumption Score (FCS), which is a proxy for household dietary diversity; and the reduced Coping Strategies Index (rCSI), which measures HH food insecurity based on their use of a number of negative coping strategies in the absence of resources (food or money). Inadequate care and feeding practices were evaluated using maternal and child health indicators including antenatal care, birth with a skilled birthing attendant, birth via C-section, childhood vaccination, care-seeking behaviors for childhood illnesses, and growth monitoring and promotion (GMP). Additionally, SBCC indicators were evaluated to assess maternal knowledge, awareness, and practices around health and IYCF. Lastly, HH environment was evaluated using indicators related to household crowding, number of CU5, access to clean water, and presence of a toilet/latrine and handwashing facilities.

Intervention: Participants were surveyed on whether they were receiving assistance or support from other nutrition-sensitive or cash programs.

Basic/Enabling Causes: To examine the basic/enabling causes of malnutrition, indicators such as household expenditure, household and personal assets and housing materials, maternal education and empowerment/decision-making, and gender of the head of household were assessed.

A detailed description of selected exposures and their definitions is included in Table S6.

### 3.3. Quantitative Tools & Equipment

Tools and software used for quantitative data were: Kobo-Toolbox (online tool development and management platform) and KoboCollect (mobile application) for data collection and storage; CommCare for quantitative data management; STATA v. 18.0 (StataCorp, College Station, TX, USA) and SAS v. 9.4 (SAS Institute Inc. Cary, NC) for quantitative analysis.

### 3.4. Quantitative Analysis

Descriptive statistics were used to summarize levels and trends in key determinants and outcomes and compared across study arms. For the main outcomes, prevalence and incidences of indicators were estimated using an intent-to-treat approach and differences were calculated between baseline, midline, and endline, and across study arms. Sensitivity analyses were also conducted, including comparison of prevalence vs. incidence approach, per protocol analysis, and complete case vs. available case analysis for handling missing data.

We also estimated the effect of the interventions on primary and secondary outcomes using mixed-effects linear regression models, controlling for baseline values and imbalances between groups. A difference-in-difference analysis design was implemented to understand statistically significant changes across timepoints and between study arms, with risk ratios estimated at a 0.05 error margin. The Bonferroni correction was applied to adjust the p-values for arms comparisons to control the family-wise error rate, and p-values<0.05 were considered statistically significant. Additionally, we interpreted and discussed the programmatic significance of the group effect sizes, given the context of our study.

## 4. Qualitative Approach

### 4.1. Focus Group Discussions – CU5 Mothers and Husbands

The focus group discussion guides (Annexes S8-9) were developed by the qualitative team in English, were distributed to the research team for feedback and suggestions and were then translated into Somali for the focus groups. A focus group discussion for each study arm (cash, cash + SBCC, cash + top-up) and in both locations (Bay and Hiran) was conducted with mothers of CU5, resulting in six (6) FGDs with mothers. One focus group in each location (Bay and Hiran), combining all 3 study arms, was conducted with the husbands, resulting in two (2) men’s FGDs. Each focus group was comprised of 8 participants, resulting in a total of 48 mothers and 16 husbands participating in the focus groups. The study team used the midline dataset as the sampling frame for the FGDs. Key criteria used to select a representative mix of participants were maternal education status, age of the mother, and whether the mother was the head of the household. For feasibility considerations, the team also considered the distance of the participant’s household from the FGD site.

Once enrollment criteria were confirmed and informed consent was obtained, trained enumerators conducted focus group discussions with mothers and husbands of CU5 in a central location in their community. Focus groups were conducted in January 2024, about one month after the endline quantitative data collection in December 2023. The goal of these discussions was to gain additional insight and perspective into the quantitative results and better understand participants’ experiences and perceptions as well as key barriers and facilitators surrounding cash for nutrition and prevention of wasting in the study area. The mother FGDs covered the following topics: the SC CashPlus for Nutrition program; the use of cash; health-seeking behaviors during pregnancy; child sickness and health-seeking behaviors when sick; and dietary diversity and food security. The same general topics were explored in the fathers’ FGDs, however, some of the questions had to be adapted to capture the husband/father’s perspective, such as exploring the topic of “supporting healthy pregnancy” instead of “health-seeking behaviors during pregnancy”.

### 4.3. Qualitative Data Analysis

All focus group discussions were recorded and transcribed. Focus group discussion transcripts in Somali were translated into English for analysis.

An iterative mix of inductive and deductive approaches was used for analysis of the focus group discussions. Since the qualitative data collection procedure followed a structured approach, the focus group discussion guides were used to inform preliminary themes and to develop initial codes. A two-phased approach was then used to develop codes with the first few available focus group discussion transcripts. The first phase included a complete readthrough of the transcript with rapid *in vivo* coding, highlighting quotes and writing down any preliminary codes. Another round of coding was then done with a higher conceptual understanding, creating additional higher-level conceptual codes. Two team members engaged in this initial coding process. A codebook detailing each code (including a brief definition, a full definition, instances of when to use and when not to use each code, and an example of text that would be coded using each code) was then developed and distributed to the qualitative data analysis team.[36] An iterative, five-phased analytic cycle approach was used for analysis, consisting of the following stages: 1) Compiling, 2) Disassembling, 3) Reassembling, 4) Interpreting, and 5) Concluding.[37] At least 2 research team members coded each focus group discussion transcript, and coded versions of each transcript were merged to create a final coded transcript for each focus group. Key informant interviews were analyzed thematically. Findings from the mothers’ focus groups, men’s focus groups, and key informant interviews were triangulated. Both exploratory analysis and explanatory sequential analysis approaches were undertaken, organizing qualitative findings according to the UNICEF conceptual framework on maternal and child nutrition and triangulating the qualitative data with the quantitative results to contextualize and further explore main study findings. After analysis of qualitative data was complete, a workshop was held with the qualitative data analysis team with members from both SC Somalia and JHU to discuss, contextualize, and interpret findings. ATLAS.ti 24 software was used for analysis of the focus group discussion transcripts. Additionally, Microsoft Office Suite was used for summarizing results and for KII analysis.

## 5. Cost-Efficiency and Cost Effectiveness Approach

The cost-efficiency and cost-effectiveness analyses were designed using the standards set by the Dioptra Systematic Cost Analysis Consortium. Analysis was conducted using a three-pronged, retrospective approach to data collection, including: 1) a desk review, 2) cost estimation, and 3) qualitative data collection from group and in-person consultations with the study implementation team. For these analyses, data was gathered regarding the program costs (financial procurement and implementation costs), project reports, results, financial reports, and survey data from the midline and endline timepoints. Cost inputs were estimated and linked to outputs and study outcomes in an iterative modelling process. Further discussions with in-country program staff were used to confirm estimates and assess intervention resource use, including all the direct, indirect, and ‘societal’ costs of the intervention.

Quantitative data was analyzed through a combination of activity-based costing (ABC) and step-down cost accounting (SDCA), where the evaluator assesses transactions and assigns costs to chosen cost categories for analysis (micro-costing). Three indicators were used to evaluate the cost efficiency of each study arm: 1) Cost per Household and 2) Cash Transfer Ratio. The cost-effectiveness analysis compared reduction in wasting prevalences and intervention costs of Arms 2 and 3 against Arm 1 (control group). Finally, the societal cost analysis was evaluated as the “opportunity cost” or time investment to beneficiary households for participating in the intervention, contributing to a holistic understanding of the intervention’s true value.

## 6. Market Monitoring

To assess the prices and availability of foods in markets throughout the study period, market monitoring activities were undertaken at markets in both Bay and Hiran (Table S10). Enumerators underwent a one-day training and then administered questionnaires to vendors in Bay and Hiran in July (48 vendors in Bay and 112 vendors in Hiran) and December 2023 (33 vendors in Bay and 78 vendors in Hiran). Additionally, the December questionnaire included recall questions for September 2023, in order to have market monitoring data corresponding with the baseline, midline, and endline data collection for the main quantitative component of the research study. Vendors were asked about the availability and prices of different commonly consumed foods and foods for an energy-based diet as well as reasons for fluctuations in price and availability. Enumerators supplemented the vendor questionnaires (Table S12) with direct observations of the markets, including taking photos, and conversations with relevant stakeholders.

Market monitoring data was analyzed in Microsoft Excel and STATA. Specific food items were categorized into one of the following domains: sentinel foods, fruits and vegetables, cereals, and protein sources (listed in Table S11). Trends in changes in food prices over time in both regions were analyzed, and vendors’ views on the reasons for fluctuations were reported.

## 7. Data Triangulation

The mixed-methods trial was designed in alignment with the trial objective and aims. Collected data was triangulated to form a holistic understanding of the study results, trial context, and participant experiences, and to inform the trial results interpretation. A figure describing the mixed-methods approach and data triangulation can be found in Figure 3 below.

**Figure 3.**
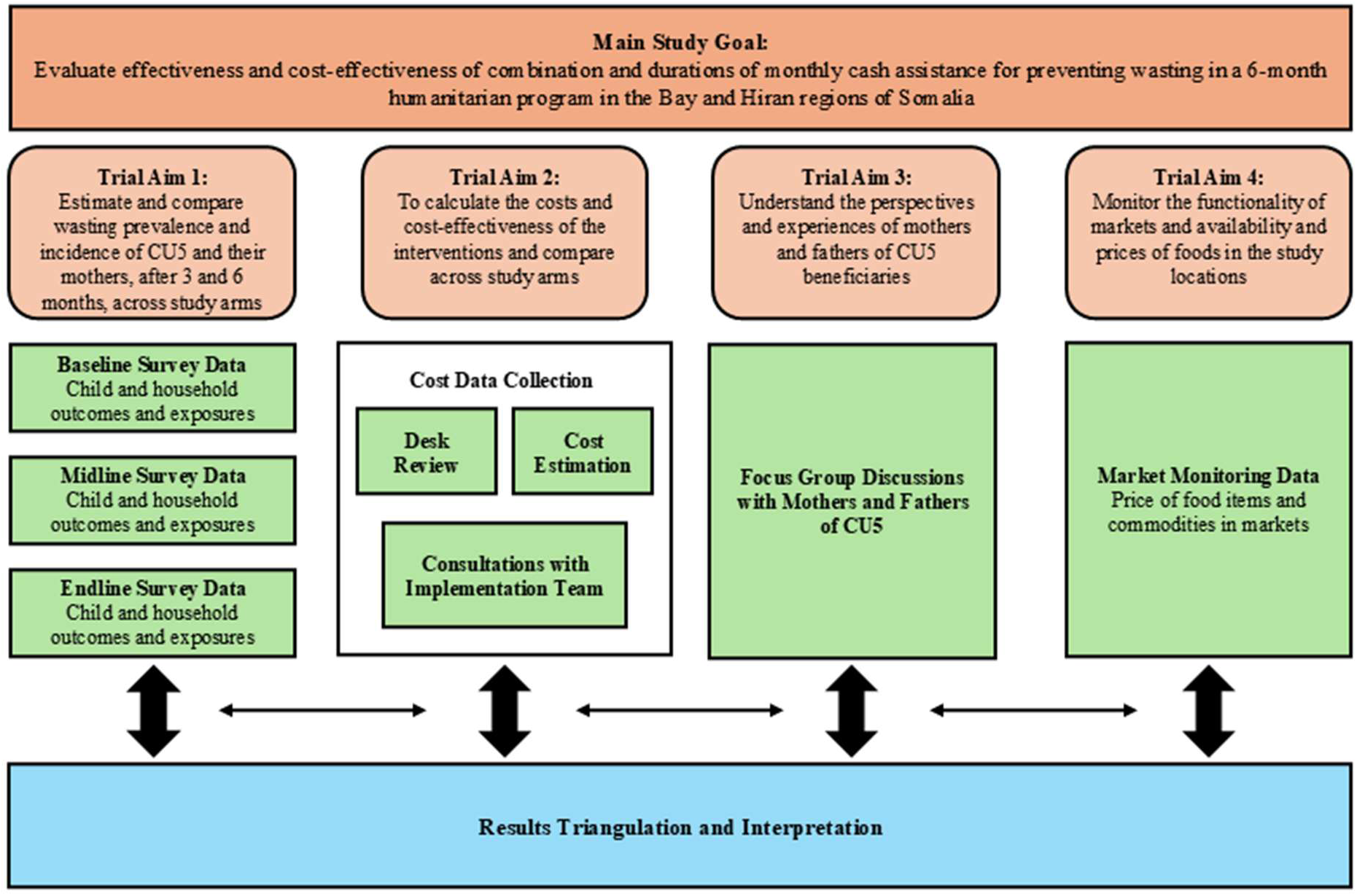
The mixed-methods approach to data collection, results triangulation and interpretation.

## 8. Adaptive Design

The approach above describes the final adaptive design used for the trial. These methods evolved throughout the design and implementation phases of the trial as new challenges and circumstances arose. A table of study adaptations may be found in Table 3 below.

**Table 3.**
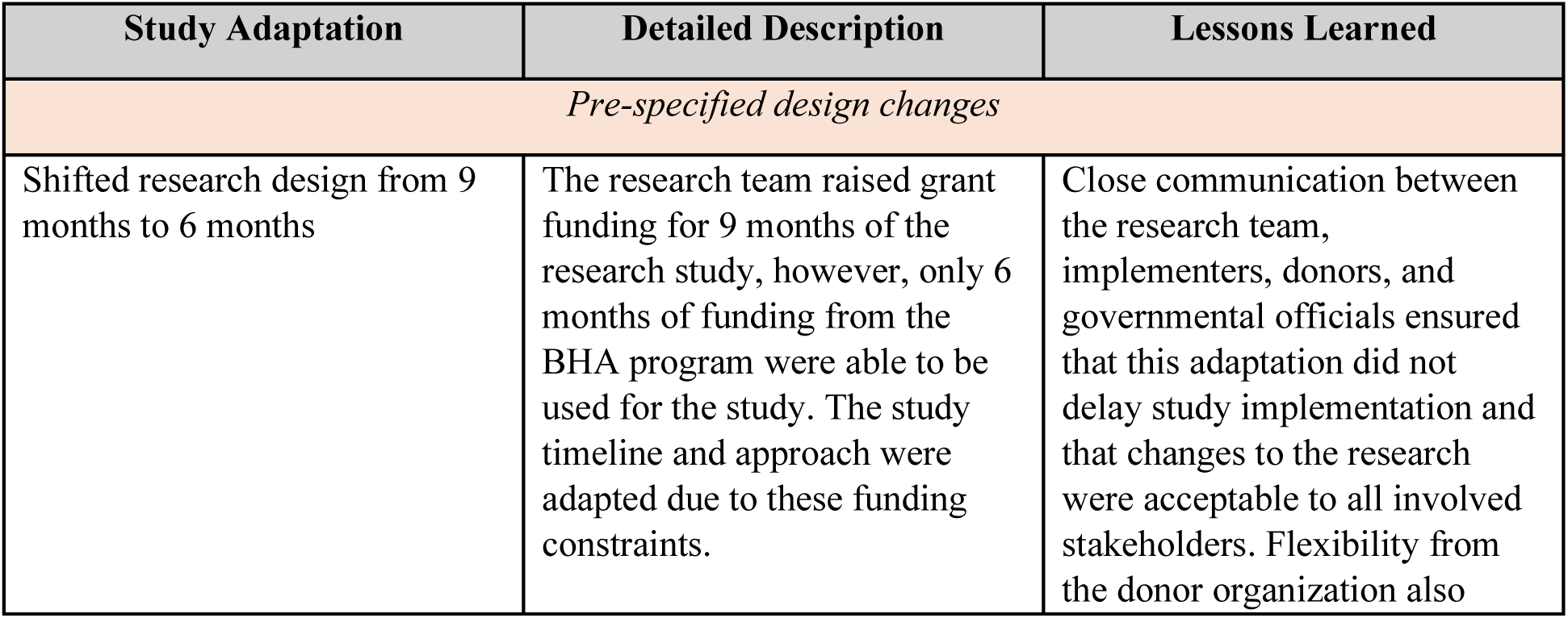
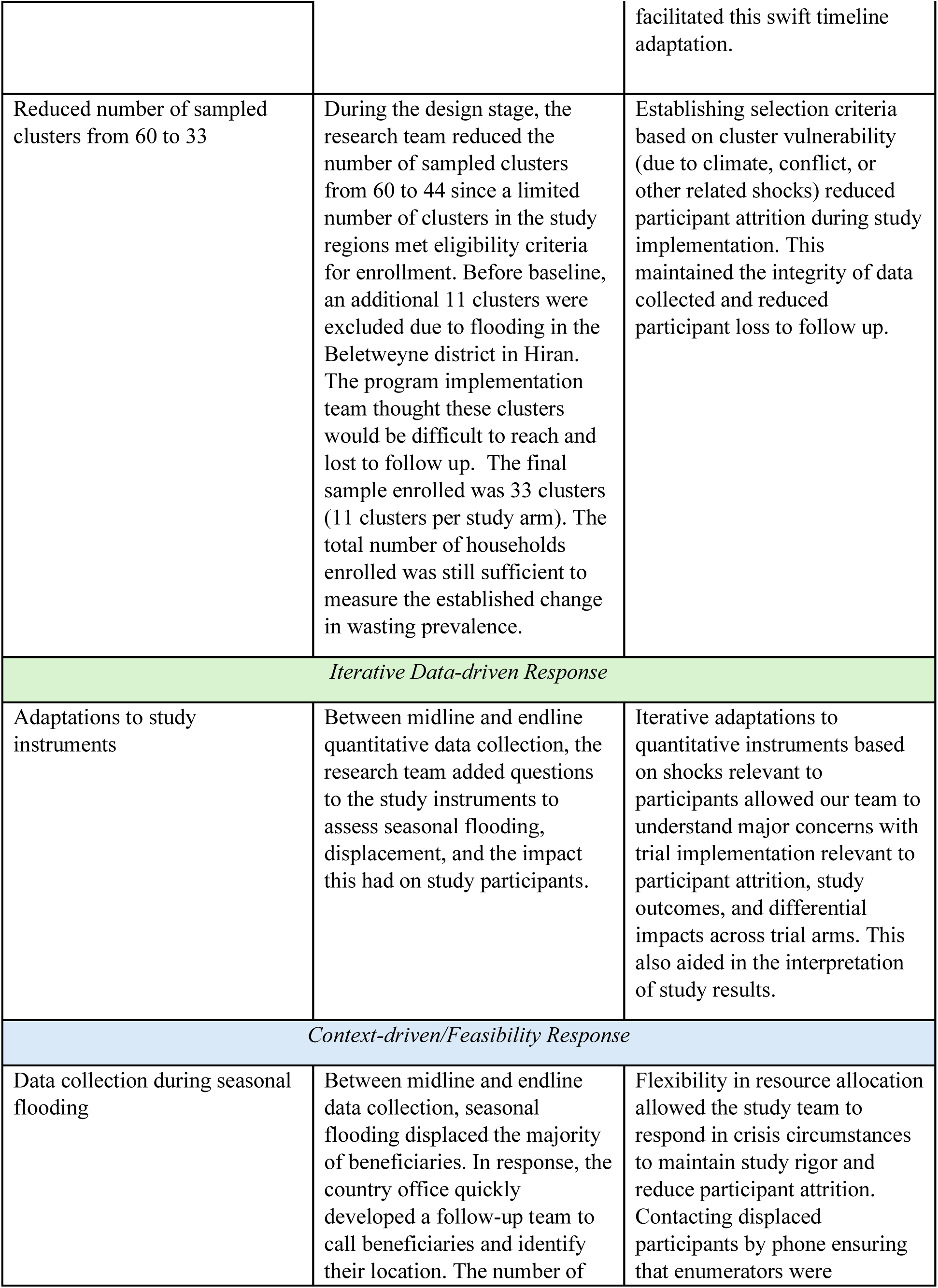
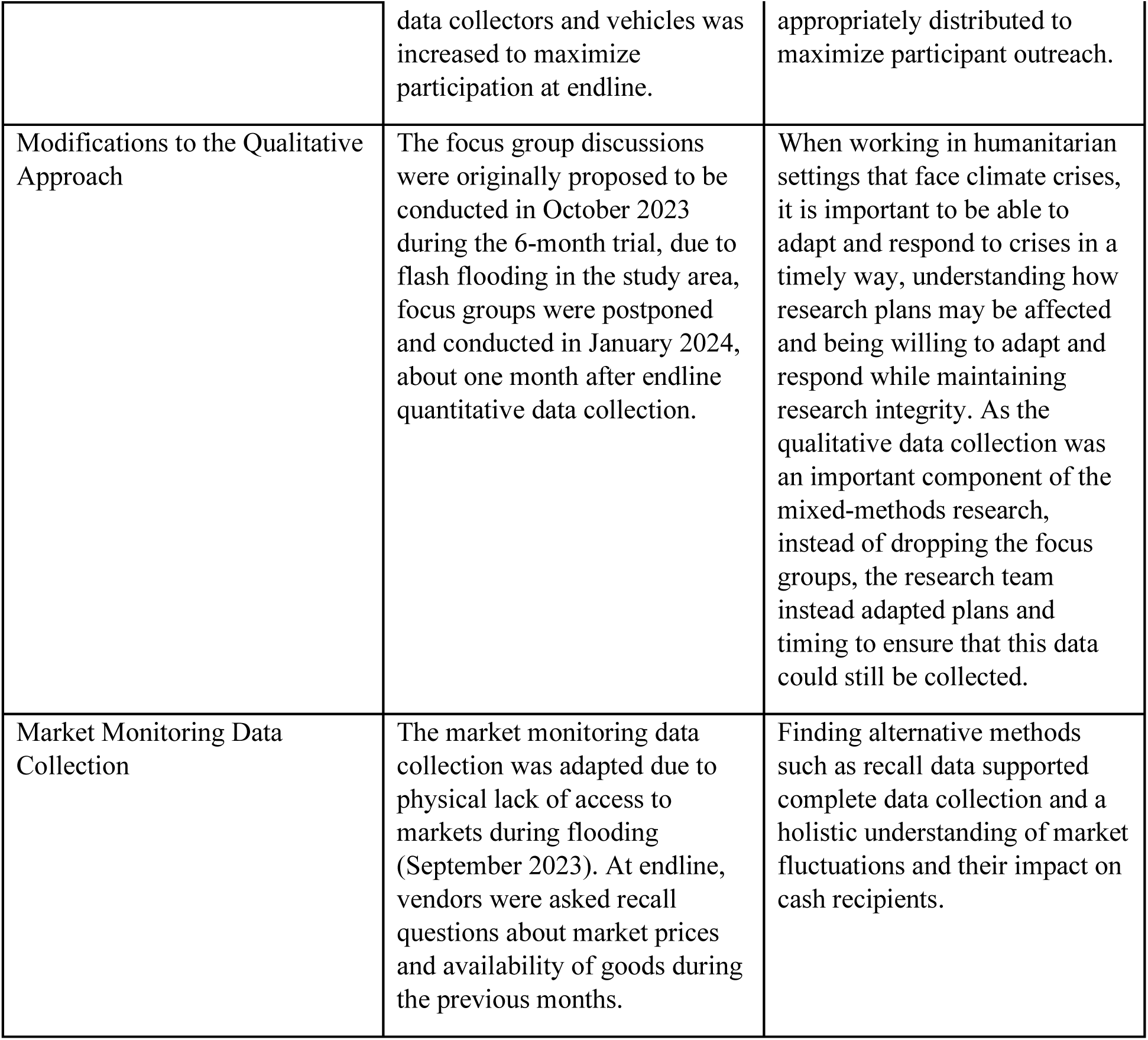
Study Adaptations.

| Study Adaptation | Detailed Description | Lessons Learned |
| --- | --- | --- |
| *Pre-specified design changes* |  |  |
| Shifted research design from 9 months to 6 months | The research team raised grant funding for 9 months of the research study, however, only 6 months of funding from the BHA program were able to be used for the study. The study timeline and approach were adapted due to these funding constraints. | Close communication between the research team, implementers, donors, and governmental officials ensured that this adaptation did not delay study implementation and that changes to the research were acceptable to all involved stakeholders. Flexibility from the donor organization also |
|  |  | facilitated this swift timeline adaptation. |
| Reduced number of sampled clusters from 60 to 33 | During the design stage, the research team reduced the number of sampled clusters from 60 to 44 since a limited number of clusters in the study regions met eligibility criteria for enrollment. Before baseline, an additional 11 clusters were excluded due to flooding in the Beletweyne district in Hiran. The program implementation team thought these clusters would be difficult to reach and lost to follow up. The final sample enrolled was 33 clusters (11 clusters per study arm). The total number of households enrolled was still sufficient to measure the established change in wasting prevalence. | Establishing selection criteria based on cluster vulnerability (due to climate, conflict, or other related shocks) reduced participant attrition during study implementation. This maintained the integrity of data collected and reduced participant loss to follow up. |
| *Iterative Data-driven Response* |  |  |
| Adaptations to study instruments | Between midline and endline quantitative data collection, the research team added questions to the study instruments to assess seasonal flooding, displacement, and the impact this had on study participants. | Iterative adaptations to quantitative instruments based on shocks relevant to participants allowed our team to understand major concerns with trial implementation relevant to participant attrition, study outcomes, and differential impacts across trial arms. This also aided in the interpretation of study results. |
| *Context-driven/Feasibility Response* |  |  |
| Data collection during seasonal flooding | Between midline and endline data collection, seasonal flooding displaced the majority of beneficiaries. In response, the country office quickly developed a follow-up team to call beneficiaries and identify their location. The number of | Flexibility in resource allocation allowed the study team to respond in crisis circumstances to maintain study rigor and reduce participant attrition. Contacting displaced participants by phone ensuring that enumerators were |
|  | data collectors and vehicles was increased to maximize participation at endline. | appropriately distributed to maximize participant outreach. |
| Modifications to the Qualitative Approach | The focus group discussions were originally proposed to be conducted in October 2023 during the 6-month trial, due to flash flooding in the study area, focus groups were postponed and conducted in January 2024, about one month after endline quantitative data collection. | When working in humanitarian settings that face climate crises, it is important to be able to adapt and respond to crises in a timely way, understanding how research plans may be affected and being willing to adapt and respond while maintaining research integrity. As the qualitative data collection was an important component of the mixed-methods research, instead of dropping the focus groups, the research team instead adapted plans and timing to ensure that this data could still be collected. |
| Market Monitoring Data Collection | The market monitoring data collection was adapted due to physical lack of access to markets during flooding (September 2023). At endline, vendors were asked recall questions about market prices and availability of goods during the previous months. | Finding alternative methods such as recall data supported complete data collection and a holistic understanding of market fluctuations and their impact on cash recipients. |

## 9. Stakeholder Engagement and Research Uptake Strategy

This methodology was co-created throughout the design and implementation of the trial through the continuous engagement of key stakeholders, which is a key feature of adaptive research methodologies.[29] This process engaged members of the research team, program staff at Save the Children, and leaders at the Ministry of Health in Somalia to ensure adaptations were acceptable and meaningful to all stakeholders involved. The methodology was co-created in an iterative process that maintained the necessary rigor and integrity of the trial while ensuring that the design would address government priorities and be practical for future researchers.

A key priority for the research team is to translate evidence into action, and the team implemented a research uptake strategy throughout the study and beyond (Annex S13).

## 10. Discussion & Lessons Learned

This trial is one of the first to apply an adaptive mixed methods design to evaluate the effectiveness and cost-effectiveness of cash interventions in the Somalia context. The adaptive design allowed us to overcome several well-documented challenges associated with conducting research in humanitarian settings, including operational constraints, partnership coordination, participant mistrust, and instability in the study environment. In this discussion, we reflect on the key factors that enabled our study to maintain rigor while addressing these challenges and offer insights for future research in similar contexts.

One of the most critical components of the trial’s success was the strong partnership coordination between the research team, program implementers, and local stakeholders. This close collaboration ensured that research methods were closely aligned with the realities of program implementation, allowing for necessary adaptations without compromising rigor. For example, when the trial faced a funding disparity—where research funding covered 9 months but cash programming was only funded for 6 months—close coordination with local staff, government officials, and donors allowed us to adjust the trial timeline and maintain our research objectives within a shortened cash program. This flexibility highlights the importance of early and continuous collaboration with on-the-ground partners in humanitarian research. Such partnerships ensure that the research remains feasible and relevant to the local context while addressing logistical and operational constraints as they arise.

The adaptive nature of the trial design was another key strength. Humanitarian settings are inherently fluid, often impacted by unpredictable events such as displacement, natural disasters, or security issues. In our study, flooding and mass displacement between midline and endline data collection posed a significant challenge, threatening both the collection of endline data and the completion of focus groups. Rather than halting the study, the adaptive design enabled the team to respond quickly. We hired local village guides to locate and mobilize displaced participants, adjusted our data collection schedule, and postponed focus groups until conditions improved. These adaptations were critical to preserving the study’s integrity and minimizing participant attrition, demonstrating that adaptive trial designs are essential for maintaining research rigor in unstable environments. This flexibility also required continuous communication with stakeholders, enabling the study to swiftly adjust its approach in response to emerging challenges.

Despite these successes, the complex environment of humanitarian research still presented several ongoing challenges. One notable issue was the difficulty in tracking participants for the societal cost analysis, particularly those who were displaced. This challenge underscores the need for future studies to incorporate more robust tracking mechanisms and data collection strategies that account for population movement, especially in regions where displacement is common.

The success of adaptive designs is often contingent upon flexible funding models and open communication with donors. Our ability to make necessary adjustments in this trial was supported by ongoing communication with our donors, which allowed for modifications to the timeline and program structure. Nonetheless, such flexibility may not always be feasible, particularly in highly regulated funding environments. Future research should advocate for funding structures that allow for adaptive designs and contingencies, recognizing that the unpredictable nature of humanitarian settings requires a departure from traditional, rigid trial frameworks.

#### Box 1. Lessons Learned

1. Early and sustained collaboration with local partners is essential to ensure that research remains feasible and contextually relevant.

2. Adaptive trial designs should be considered a valuable tool for navigating the dynamic nature of humanitarian contexts.

3. Flexibility must extend beyond the research team to include donors and funding agencies. Future research initiatives should advocate for funding models that allow for adaptive timelines and emergency contingencies to ensure studies can continue, even under evolving conditions.

## 11. Conclusion

This study presents the mixed-methods approach used in a trial evaluating the effectiveness and cost-effectiveness of cash assistance interventions for reducing child and maternal wasting in two regions of Somalia. The **adaptive methodology**, **flexibility and responsiveness**, and **strong partnerships** between all relevant stakeholders were critical to the trial’s success. These elements ensured that the study could navigate the complex challenges of the humanitarian context, including operational constraints and population displacement, while maintaining methodological rigor. By incorporating flexibility in both research design and funding models, future studies will be better equipped to handle unforeseen challenges while producing reliable and actionable findings. In conclusion, the methodology and experiences shared here will contribute to more responsive and resilient research in humanitarian settings, ensuring that future studies are both rigorous and aligned with the needs of those most affected by crises.

## Author Contributions

Conceptualization of the original study protocol proposal and methodology: SW, QK, MT, FL, SAM, MAN, MBM, DIJ, DG, MO, MO, AA, AAM, AA, NA; writing (original draft preparation) – SW, KKA, SG, SG, QK, EM, NA; writing (review and editing) – SW, KKA, SG, SG, QK, MT, FL, SAM, MAN, SA, EM, MO, MO, AA, AAM, AA, NA. All authors have read and agreed to the published version of the manuscript.

## Institutional Review Board Statement

The study was conducted in accordance with the Declaration of Helsinki and approved by the Institutional Review Board of the Johns Hopkins Bloomberg School of Public Health (protocol code 24476, date of approval: May 17, 2023), the Ministry of Health and Human Services in Somalia (XAG/203/23, date of approval: April 26th, 2023), and the Save the Children International Ethics Review Committee (ERC).

## Data Availability

This data was collected on a vulnerable population linked to a large-scale cash assistance program in Somalia. The data may be available upon request with permission from local study investigators.

## Informed Consent Statement

Informed consent was obtained from all subjects involved in the study.

## Acknowledgments

We acknowledge the important contributions of all study participants and their families; the Ministries of Health in Somalia; and the teams of dedicated staff who collected data.

## References

1 Integrated Food Security Phase Classification. Somalia: Acute Food Insecurity Situation January - March 2023 and Projection for April - June 2023. 2023 Feb. Available: https://www.ipcinfo.org/ipc-country-analysis/details-map/en/c/1156238/. Accessed: 20 February 2025.

2 International Organization for Migration. Somalia Crisis Response Plan 2024. 2024 Mar. Available: https://crisisresponse.iom.int/response/somalia-crisis-response-plan-2024. Accessed: 20 February 2025.

3 Food Security and Nutrition Analysis Unit. Somalia 2023 Post Gu Acute Malnutrition Median Prevalence by District (Estimated). Food and Agriculture Organization of the United Nations. 2023. Available: https://fsnau.org/downloads/somalia-2023-post-gu-total-acute-malnutrition-burden-district-21-sep-2023. Accessed: 20 February 2025.

4 Blanchet K, Sistenich V, Ramesh A, Frison S, Warren E, Smith J, et al. An evidence review of research on health interventions in humanitarian crises. 2015 Oct. Available: https://www.elrha.org/wp-content/uploads/2015/01/Evidence-Review-22.10.15.pdf. Accessed: 20 February 2025.

5 Doocy S, Lyles E, Tappis H. An evidence review of research on health interventions in humanitarian crises: 2021 Update. 2022 Jun. Available: https://www.elrha.org/wp-content/uploads/2022/06/Elrha-HHER2-FullReport.pdf. Accessed: 20 February 2025.

6 Woodward A, Griekspoor A, Doocy S, Spiegel P, Savage K. Research agenda-setting on cash programming for health and nutrition in humanitarian settings. Journal of International Humanitarian Action. 2018;3:7. doi:10.1186/s41018-018-0035-6.

7 Durr A, Cluster GN, UNICEF, CashCap/NORCAP. Evidence and Guidance Note on the Use of Cash and Voucher Assistance for Nutrition Outcomes in Emergencies. 2020 Aug. Available: https://www.nutritioncluster.net/sites/nutritioncluster.com/files/2021-02/UNICEF_Cash-report_EN_RGB_170221_V9_FINAL_0.pdf. Accessed: 20 February 2025.

8 Yavuz C, Villar PF, Berretta M, Nabi A, Cooper C, Shisler S. Improving Food Security in Humanitarian Emergencies: An Evidence Gap Map. 2022. Available: https://fsnnetwork.org/sites/default/files/2023-02/Improving_food%20security_in_humanitarian_emergencies_An_evidence_gap_map_HAEC_0.pdf. Accessed: 20 February 2025.

9 VanRooyen M. The Need for Humanitarian Research: Addressing Emerging Challenges in a Complex World. Harvard Humanitarian Initiative. 2024. Available: https://hhi.harvard.edu/news/need-humanitarian-research-addressing-emerging-challenges-complex-world. Accessed: 20 February 2025.

10 Smith J, Blanchet K. Research Methodologies in Humanitarian Crises. 2020. Available: https://www.elrha.org/wp-content/uploads/2020/02/R2HC-Research-Methodologies-in-Humanitarian-Crises-new.pdf. Accessed: 20 February 2025.

11 Kohrt BA, Mistry AS, Anand N, Beecroft B, Nuwayhid I. Health research in humanitarian crises: an urgent global imperative. BMJ Glob Health. 2019;4:e001870. doi:10.1136/bmjgh-2019-001870.

12 Blanchet K, Ramesh A, Frison S, Warren E, Hossain M, Smith J, et al. Evidence on public health interventions in humanitarian crises. The Lancet. 2017;390:2287–96. doi:10.1016/S0140-6736(16)30768-1.

13 Ben-Eltriki M, Rafiq A, Paul A, Prabhu D, Afolabi MOS, Baslhaw R, et al. Adaptive designs in clinical trials: a systematic review-part I. BMC Med Res Methodol. 2024;24:229. doi:10.1186/s12874-024-02272-9.

14 Odjidja EN, dos Reis AA, Engagement GC, Fund R. Learning from adaptive evaluation approaches in dynamic conflict-afflicted and humanitarian settings. African Development Bank Evaluation Matters Magazine: Learning from Successes and Failures in Evaluation. 2021. Available: https://idev.afdb.org/en/content/learning-adaptive-evaluation-approaches-dynamic-conflict-afflicted-and-humanitarian. Accessed: 20 February 2025.

15 Bhandari N, Upadhyay RP, Chowdhury R, Taneja S. Challenges of adopting new trial designs in LMICs. Lancet Glob Health. 2021;9:e575–6. doi:10.1016/S2214-109X(21)00168-6.

16 Integrated Food Security Phase Classification (IPC). IPC Acute Food Insecurity and Acute Malnutrition Analysis: Somalia. Integrated Food Security Phase Classification. 2023. Available: https://www.ipcinfo.org/ipc-country-analysis/details-map/en/c/1156238/. Accessed: 20 February 2025.

17 European Union Agency for Asylum. Country Guidance: Somalia: Hiraan. 2022 Jun. Available: https://euaa.europa.eu/country-guidance-somalia-2022/hiraan. Accessed: 20 February 2025.

18 European Union Agency for Asylum. Country Guidance: Somalia: Bay. 2022 Jun. Available: https://euaa.europa.eu/country-guidance-somalia-2022/bay. Accessed: 20 February 2025.

19 City Population, Brinkhoff T. Hiiraan. 2021. Available: https://www.citypopulation.de/en/somalia/admin/20hiiraan/. Accessed: 20 February 2025.

20 City Population, Brinkhoff T. Bay. 2021. Available: https://www.citypopulation.de/en/somalia/admin/24bay/. Accessed: 20 February 2025.

21 Thalheimer L, Schwarz MP, Pretis F. Large weather and conflict effects on internal displacement in Somalia with little evidence of feedback onto conflict. Global Environmental Change. 2023;79:102641. doi:10.1016/j.gloenvcha.2023.102641

22 ACAPS. Somalia: Key Crises to watch in 2023. 2023 Apr. Available: https://www.acaps.org/fileadmin/Data_Product/Main_media/20230413_acaps_thematic_report_somalia_key_crises_to_watch_in_2023.pdf. Accessed: 20 February 2025.

23 ACAPS. ACAPS Briefing Note: Somalia - Flooding in Baidoa. 2023 Oct. Available: https://www.acaps.org/fileadmin/Data_Product/Main_media/20231026_ACAPS_Briefing_Note_Somalia_flooding_in_Baidoa.pdf. Accessed: 20 February 2025.

24 United Nations Office for the Coordination of Humanitarian Assistance Somalia. Somalia: 2023 Deyr Season Floods Bi-Weekly Situation Report No. 5 (As of December 2023). 2023 Dec. Available: https://reliefweb.int/report/somalia/somalia-2023-deyr-season-floods-situation-report-no-5-24-december-2023. Accessed: 20 February 2025.

25 The United Nations Refugee Agency, UNHCR, Norwegian Refugee Council. Somalia Internal Displacement. 2017 [cited 16 Feb 2025]. Available: https://unhcr.github.io/dataviz-somalia-prmn/index.html. Accessed: 20 February 2025.

26 European Union Agency for Asylum. Country Guidance: Somalia (August 2023). 2023 Aug. Available: https://euaa.europa.eu/publications/country-guidance-somalia-august-2023. Accessed: 20 February 2025.

27 United Nations Children’s Fund. UNICEF Conceptual Framework on Maternal and Child Nutrition, 2020. 2021 Nov. Available: https://www.unicef.org/documents/conceptual-framework-nutrition. Accessed: 20 February 2025.

28 United Nations Children’s Fund. Improving Child Nutrition: The Achievable Imperative for Global Progress. UNICEF. 2013. Available: https://data.unicef.org/resources/improving-child-nutrition-the-achievable-imperative-for-global-progress/. Accessed: 20 February 2025.

29 Gokhale S, Walton M. Adaptive Evaluation: A Complexity-Based Approach to Systematic Learning for Innovation and Scaling in Development. 2022 Sep. Available: https://www.hks.harvard.edu/centers/cid/publications/faculty-working-papers/adaptive-evaluation-complexity-based-approach-systematic-learning-innovation-and-scaling. Accessed: 20 February 2025.

30 National Bureau of Statistics, Federal Republic of Somalia. 2022 Somalia Integrated Household Budget Survey. 2023 Feb. Available: https://nbs.gov.so/wp-content/uploads/2024/01/SOMALIA-INTEGRATED-HOUSEHOLD-BUDGET-SURVEY-SIHBS-2022.pdf. Accessed: 20 February 2025.

31 Grijalva-Eternod C, Jelle M, Haghparast-Bidgoli H, Colbourn T, Golden K, King S, et al. A cash-based intervention and the risk of acute malnutrition in children aged 6–59 months living in internally displaced persons camps in Mogadishu, Somalia: A non-randomised cluster trial. PLoS Med. 2018;15:e1002684. Available: 10.1371/journal.pmed.1002684.

32 Seidenfeld D, Handa S, de Hoop T, Morey M. Intraclass Correlations Values in International Development: Evidence Across Commonly Studied Domains in sub-Saharan Africa. Eval Rev. 2023;47:786–819. Medline:36729038 doi:10.1177/0193841X231154714.

33 Standardized Monitoring and Assessment of Relief and Transitions (SMART). Measuring mortality, nutritional status, and food security in crisis situations, Version 1. 2006 Aug. Available: http://www.nutrisurvey.de/ena_beta/SMART_Methodology_08-07-2006.pdf. Accessed: 20 February 2025.

34 World Health Organization. Guideline: updates on the management of severe acute malnutrition in infants and children. Geneva: World Health Organization; 2013. Available: https://www.ncbi.nlm.nih.gov/books/NBK190328/.

35 Tang AM, Chung M, Dong KR, Bahwere P, Bose K, Chakraborty R, et al. Determining a global mid-upper arm circumference cut-off to assess underweight in adults (men and non-pregnant women). Public Health Nutr. 2020;23:3104–13. Medline:32799964 doi:10.1017/S1368980020000397.

36 Guest G, Macqueen K. Handbook for Team-Based Qualitative Research. AltaMira Press; 2008.

37 Yin RK. Qualitative Research from Start to Finish. 2nd ed. New York: Guilford Press; 2015.

