## Supplemental Files for "A Cluster-Randomized Trial to Compare Effectiveness and Cost-effectiveness of Cash Plus Interventions in Preventing Child Wasting in Somalia: An Evidence-Based Methodology"

Methodology Paper Supplemental Materials S1-S14

**Methodology Paper Supplement Materials - Table of Contents**

Figure S1. UNICEF Conceptual Framework on the Determinants of Maternal and Child Nutrition, 2020

Annex S2. Explanation of SBCC Components

Annex S3. Table of Subject Recruitment

Table S4. Baseline Survey Instrument

Table S5. Midline Survey Instrument

Table S6. Endline Survey Instrument

Table S7. Exposures: Household Survey Indicators and Definitions

Annex S8. Mothers Focus Group Discussion Guide

Annex S9. Men's Focus Group Discussion Guide

Table S10. Market Monitoring Data Collection Sites

Table S11. Food Commodities Monitored During Market Monitoring

Table S12. Market Monitoring Questionnaire

Annex S13. Research Uptake Strategy

Table S14. Mobile Messaging Received by BHA Participants

**Figure S1. UNICEF Conceptual Framework on the Determinants of Maternal and Child Nutrition, 2020**

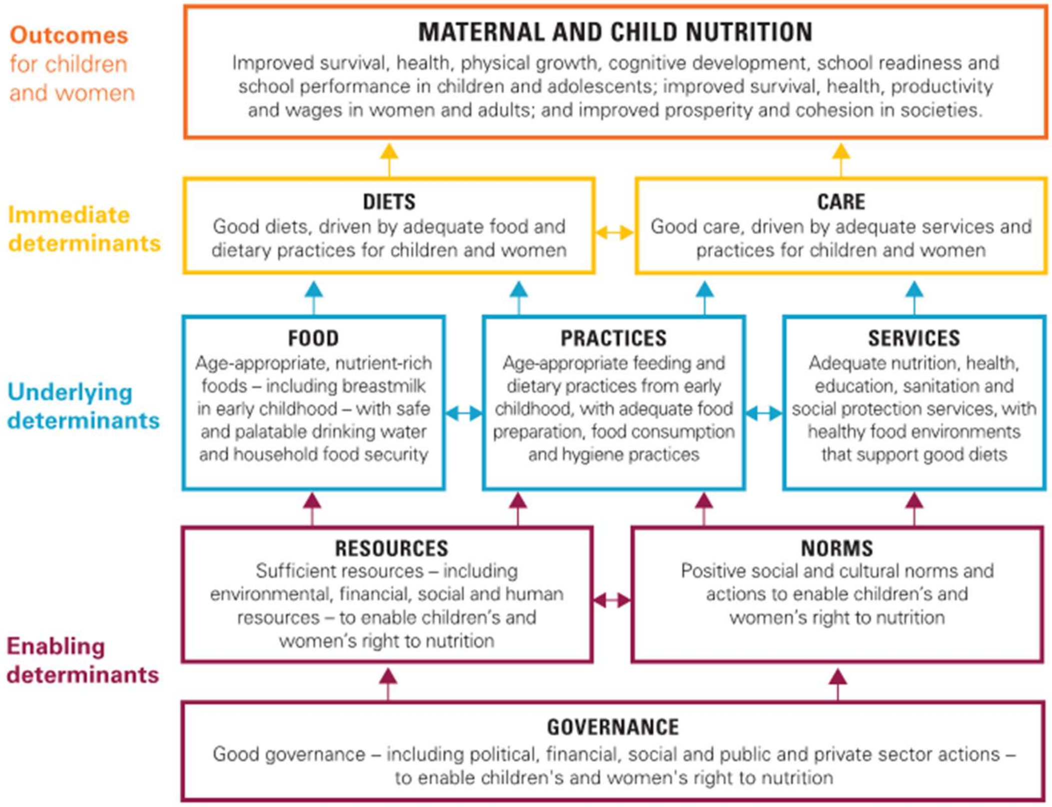

**UNICEF Conceptual Framework on the Determinants of Maternal and Child Nutrition, 2020.**
**A framework for the prevention of malnutrition in all its forms.**

**Annex S2. Explanation of SBCC Intervention Component**

SBCC sessions covered the following topics: breastfeeding within the first hour/importance of colostrum; exclusive breastfeeding for 6 months; attachment and positioning of the baby at the breast; breastfeeding misconceptions and myths; complementary feeding (continuing breastfeeding and giving other foods/liquids); medical care; hygiene; common conditions that can affect breastfeeding; physical and mental stimulation of the child; relationships (i) with partner/husband and (ii) with other family members, (e.g. mother-in-law); mother's safety/fear/worries; nutrition of pregnant and lactating mothers.

Save the Children identified 11 staff members to serve as community-based IYCF promoters to train mothers in IYCF practices and engagement strategies for mother-to-mother (M2M) support groups (4 promoters in Bay and 7 in Hiran). SBCC M2M sessions were then led by trained lead mothers who facilitated discussions among 10-15 mothers. Sessions occurred weekly, covering one of the above topics each week (for a total of 12 sessions in 12 weeks), and group members decided on the day of the sessions. Sessions lasted from 45-60 minutes. The SBCC sessions were held in a safe place to enable mothers to exchange ideas, share experiences, give and receive information, and offer and receive support in breastfeeding, child-rearing, and women's health. The lead mother also followed-up with and managed absenteeism during sessions.

In addition to the SBCC sessions, Save the Children organized cooking/food demonstrations for support groups to showcase locally available and affordable nutritious food.

Some mothers enrolled in Arm 2 missed some of the Mother-to-Mother Support Group (MtMSG) sessions, while others migrated due to search of pasture and water for their livestock. The implementation team provided make-up sessions for the mothers who missed some MtMSG sessions. For those who migrated, the implementation team attempted to identify their new locations and collaborated with the local community to arrange MtMSG sessions that were more accessible.

**Table S3. Table of Subject Recruitment**

| Subject | Data Collection | Frequency | Location and Amount of Time per Encounter | Total # of Participants | Per Arm (1 of 3 research arms) |
|---|---|---|---|---|---|
| Beneficiary child (CU5) | Growth measurements (MUAC, Weight, Height) | Baseline, Midline, Endline | Beneficiary home, up to 20 minutes | 1,894 | Arm 1: 672<br>Arm 2: 653<br>Arm 3: 569 |
| CU5 Mothers | Growth measurements (MUAC) | Baseline, Midline, Endline | Beneficiary home, 5 minutes | 1,490 | Arm 1: 511<br>Arm 2: 507<br>Arm 3: 472 |
| CU5 Mothers | Household Interview | Baseline, Midline, Endline | Beneficiary home, 45 minutes | 1,490 | Arm 1: 511<br>Arm 2: 507<br>Arm 3: 472 |
| CU5 Mothers | Focus Group Discussion | After Endline | Accessible community location, 60 minutes | 48 | 16 per arm |
| Husbands of CU5 Mothers | Focus Group Discussion | After Endline | Accessible community location, 60 minutes | 16 | N/A |
| Cash/Nutrition Stakeholders, Global | Key Informant Interviews | After Endline | Virtual, 30-60 minutes | 3 | N/A |
| **Total** | | | | **3,451 (total# of unique participants)** | |

**Table S4. Baseline Survey Instrument**

| **Baseline Questionnaire** |
|---|
| **Enumerator and Location Details** |
| Start Time |
| End Time |
| Date |
| Enumerator #1 ID |
| Enumerator #2 ID |
| Enumerator #3 ID |
| Team ID |
| Household Unique ID |
| Household Member Unique ID (cash recipient) |
| Unique ID |
| Displacement Status |
| Region |
| District |
| Village |
| Cluster Number |
| Study Arm |
| **Informed Consent** |
| Hello, we are from Save the Children, and we would like to speak with {mother's name}. Can we talk to you? |
| Thank you for the opportunity to speak with you. As I mentioned, we are from Save the Children. We are conducting a survey with randomly selected program households to learn more about the effect of our multi-purpose cash program. The questions will take about **45 minutes** to complete.<br><br>Your participation is voluntary. If you agree to participate, you can choose to stop at any time or skip any questions you do not want to answer. There will be no compensation for participating in this survey. If you do not agree to participate, it will not affect your status in the program or the services you receive.<br><br>Any personal information such as names that you share will be kept confidential and only a limited number of Save the Children staff will have access to it. We intend to share the resulting analysis, which will not include personal information, with our donors and partners to help improve future programs. Let me know if you have any questions about this.<br><br>If you have additional questions after this, you can also contact [MEAL team must insert pre-selected staff member name and phone number into form before deployment]. |
| Do you agree and consent to be interviewed? |
| Enumerator Name (acknowledging that consent has been obtained) |
| Enumerator Signature (acknowledging that consent has been obtained) |
| Thank you for your time. |

| **Respondent Information** |
|---|
| Mother's Name |
| Mother's Age |
| What is the highest level of school you attended, including madrasa? |
| Are you currently pregnant? |
| Are you the head of the household? |
| If no, what is the gender of the head of household? |
| What is your relationship with the head of the household? |
| Who usually decides how the household income/earnings (all sources of income) will be used: you, your husband, or you and your husband jointly? |
| If other, please specify. |
| Who usually makes decisions about healthcare for yourself: you, your husband, or you and your husband jointly? |
| If other, please specify. |
| In the last 12 months, has any member of your household received **any assistance from any institution** such as the government, international organizations, religious bodies in the form of...?<br>None<br>Cash<br>Food, including school feeding<br>Other in-kind, such as animals<br>Micronutrient supplementation (vitamin A, iron, etc)<br>Specialized Nutritious Food (SNF)<br>WASH (hygiene, water, etc)<br>Scholarship<br>Other (please specify) |
| If other, please specify. |
| How much did you receive (either total value in USD or kg or other unit)? |
| How often did you receive it? (weekly, monthly, etc.) |
| **Child Anthropometrics** |
| How many children under 5 years of age are currently living in this household? |
| Name of the child |
| Sex of the child |
| What is your relationship to this child? |
| Age of the child (in months) |
| How did you confirm the age?<br>Seasonal Calendar<br>Health card<br>Birth certificate |
| Child’s Weight to the nearest 0.1kg |
| Child’s Height to the nearest 0.1cm |
| Child’s MUAC to the nearest 0.1cm |
| Is oedema present in the child? |
| How many children under 5 years of age are currently living in this household? |

| |
|---|
| Enumerator only: record the members of the household that are present during the interview (such as the mother, head of household, children, other family members, etc.) |
| **Maternal Wasting: MUAC** |
| Mother MUAC |
| Weight (kg) to the nearest 0.1kg |
| **Household Expenditures** |
| For the next set of questions I am going to ask you to estimate how much your household spent **in USD** on different household needs over the past month (30 days). |
| How much would you estimate you spent in total on **all your food** over the last 7 days? |
| How much would you estimate you spent in total on **nutritious food** over the last 7 days? |
| Besides food, what are the other 3 main expenses your household spends the most on each month?<br>Hygiene items (soap, toothbrush, toothpaste, toilet paper, etc.)<br>Utilities (electricity, water, etc.)<br>Healthcare (doctor's visits, medicine, child vaccinations, ante-natal or post-natal care)<br>Household NFIs (kitchen items, clothing, shoes, etc.)<br>Education (tuition, school supplies, uniforms)<br>Debt repayment |
| In the last month (30 days) how much did your household spend on hygiene products (soap, toothbrush, toothpaste, toilet paper, etc.)? |
| In the last month (30 days) how much did your household spend on utilities (water, electricity, rent)? |
| In the last month (30 days) how much did your household spend on medical costs like doctor's visits and medicine, ante- or post-natal care, child vaccinations? |
| In the last month (30 days) how much did your household spend on non-food items like clothing, shoes, kitchen supplies? |
| In the last month (30 days) how much did your household spend on school fees, school supplies, books, school uniforms, and other education expenses? |
| In the last month (30 days) how much did your household spend on debt repayment? |
| **Underlying Causes – Household Food Insecurity** |
| **Food Consumption Score** |
| I would like to ask you about all the different foods that your household members have eaten in the last 7 days. Could you please tell me how many days in the past week your household has eaten the following foods? |
| In the past week, approximately how many days has your household eaten cereals and tubers such as maize, maize porridge, rice, sorghum, millet, pasta, bread and other cereals, cassava/yucca, potato, or sweet potato? |
| Yes/No? |
| Number of Days |
| In the past week, approximately how many days has your household eaten legumes and nuts such as beans, peas, groundnuts, or cashew nuts? |
| Yes/No? |
| Number of Days |
| In the past week, approximately how many days has your household eaten vegetables or leaves? |

| |
|---|
| Yes/No? |
| Number of Days |
| In the past week, approximately how many days has your household eaten fresh fruits? |
| Yes/No? |
| Number of Days |
| In the past week, approximately how many days has your household eaten eggs or meat such as beef, goat, poultry, camel, or fish? |
| Yes/No? |
| Number of Days |
| In the past week, approximately how many days has your household eaten dairy products such as milk, yogurt, or cheese? |
| Yes/No? |
| Number of Days |
| In the past week, approximately how many days has your household eaten sugar and sugary products such as honey, jam, candy, or pastries. |
| Yes/No? |
| Number of Days |
| In the past week, approximately how many days has your household eaten oils, fats or butter? |
| Yes/No? |
| Number of Days |
| In the past week, approximately how many days has your household eaten condiments such as spices, tea, coffee, salt, fish powder, or small amounts of milk for tea? |
| Yes/No? |
| Number of Days |
| Total FCS Score |
| Category FCS |
| IPC Phase FCS |
| FCS: High Oil |
| IPC Phase FCS: High Oil |
| **Reduced Coping Strategies Index (rCSI)** |
| In the past 7 days, if there have been times when you did not have enough food or money to buy food, how many days has your household had to: |
| a. Rely on less preferred and less expensive foods? |
| Yes/No? |
| Number of Days |
| b. Borrow food, or rely on help from a friend or relative? |
| Yes/No? |
| Number of Days |
| c. Limit portion size at mealtimes? |
| Yes/No? |
| Number of Days |
| d. Restrict consumption by adults in order for small children to eat? |

| |
|---|
| Yes/No? |
| Number of Days |
| e. Reduce number of meals eaten in a day? |
| Yes/No? |
| Number of Days |
| Total RCSI Score |
| IPC Phase RCSI Score |
| **Household Hunger Scale** |
| In the past 30 days, was there ever no food to eat of any kind in your house because of lack of resources to get food?<br>How often did this happen in the past 30 days? |
| In the past 30 days, did you or any household member go to sleep at night hungry because there was not enough food?<br>How often did this happen in the past 30 days? |
| In the past 30 days, did you or any household member go a whole day and night without eating anything at all because there was not enough food?<br>How often did this happen in the past 30 days? |
| HHS Score Total |
| HHS Category |
| HHS IPC Phase |
| **Underlying Causes – Unhealthy HH environment** |
| What is the main source of drinking water for members of your household?<br>If other, specify. |
| Does the household have access to a handwashing place that has soap and water?<br>What type of handwashing station is it?<br>Sink<br>Bathroom<br>Laundry<br>Other, please specify |
| Is water present for handwashing? |
| Is cleansing agent available for handwashing? |
| When do wash your hands (after what type of daily activities)?<br>Before preparing food<br>Before eating<br>Before feeding children<br>After handling a child's stool/diaper<br>After using latrine/toilet<br>Other (specify) |
| When do you think are the three (3) most important times to wash hands?<br>Before preparing food<br>Before eating<br>Before feeding children<br>After handling a child's stool/diaper<br>After using latrine/toilet |

| |
|---|
| Other (specify) |
| What kind of toilet facility do members of your household usually use? |
| If other, specify. |
| **Underlying Causes – Inadequate Care & Feeding Practices** |
| What is the age in months of the youngest child living this household? |
| Thinking about your youngest child, can you please respond to the following questions about health care services. |
| What is the name of your youngest child |
| Did you see anyone for antenatal care for this pregnancy?<br> Why did you not see anyone for antenatal care?<br> Who did you see?<br> If other, please specify. |
| How many times did you received care during this pregnancy? |
| During your last pregnancy, did you take any any tablets or syrup that contain iron? |
| During your last pregnancy did you receive any kind of food assistance for yourself?<br> What kind of foods did you receive?<br> Ready-to-use food (RUSF, Plumpy' product, Lipid-based nutrient supplements (packaged pastes))<br> Special flour (Corn Soy Blend/CSB, Super Cereal/SC)<br> High energy Biscuit<br> Food basket with mix of products (rice, oil, beans, wheat)<br> Fortified beverage<br> Other, specify<br> No response |
| Who assisted with the delivery of your youngest child?<br> If other, please specify. |
| Where did you give birth to your youngest child?<br> If other, please specify. |
| Was your youngest child delivered by caesarean (did they cut your belly open to take the baby out)? |
| When your youngest child was born, was your youngest child very large, larger than average, average, smaller than average, or very small? |
| When you were pregnant with your youngest child, did you have any illness that required you to seek health care or treatment beyond your regular pregnancy check-ups?<br> If yes, what were the illnesses?<br> Diabetes<br> High blood pressure<br> Other |
| **Child Vaccination History** |
| Thinking about your youngest child, your youngest child, did s/he ever receive any vaccinations to prevent diseases, including vaccinations received in campaigns or immunization days or child health days? |

| |
|---|
| Has your youngest child ever received any of the following vaccinations?<br>BCG against TB (injection in the arm/shoulder that usually causes a scar)<br>Oral polio vaccine (about 2 drops in the mouth)<br>Pentavalent (injection in the thigh sometimes at the same time as polio drops)<br>Measles vaccination (injection in the arm) |
| How many times did your youngest child receive the pentavalent vaccine? |
| How many times did your youngest child receive the measles vaccine? |
| **Child Health & Nutrition** |
| Has your youngest child had diarrhea, fever, or cough in the last 2 weeks?<br>None<br>Diarrhea<br>Ill with a fever<br>Ill with a cough |
| Did you seek treatment for the illness from any source? |
| **Maternal and Child Wasting** |
| In the last 3 months, did you and your youngest child have your and their height, weight or arm circumference measured in the facility, in the community or in your home? |
| In the last 6 months, were you and your youngest child sent for treatment after the height, weight or arm circumference were measured? |
| If yes, did you or your youngest child receive the treatment? |
| If yes, what did they receive during the treatment? For example, CSB++ (porridge), Plumpy Nut or medication or anything else?<br>CSB++ (porridge)<br>Plumpy Nut<br>Other, please specify |
| **Immediate Causes - Inadequate Dietary Intake** |
| Minimum Dietary Diversity for Children (MDD-C) |
| Has your youngest child ever been breastfed? |
| How long after birth did you put your youngest child to the breast for the first time?<br>Hours?<br>Days? |
| Was your youngest child breastfed yesterday during the day or at night? This includes any breast milk the child drank from a bottle, cup, or spoon as well as directly from the breast.<br>If yes, how many times did you breastfeed your child? |
| Now I would like to ask you about some medicines and vitamins that are sometimes given to infants.<br>Was your youngest child given any vitamin drops or other medicines as drops yesterday during the day or at night? |
| Was your youngest child given oral rehydration solution yesterday during the day or at night? |

| |
|---|
| Next, I would like to ask you about some liquids that your youngest child may have had yesterday during the day or at night. Did your youngest child have any of the following…<br>Plain water?<br>Infant formula?<br>How many times yesterday during the day or at night did your youngest child consume any formula?<br>Any milk such as tinned, powdered, or fresh animal milk?<br>How many times yesterday during the day or at night did your youngest child consume any of these types of milk?<br>Any juice or juice drinks?<br>Clear broth?<br>Yogurt?<br>How many times yesterday during the day or at night did your youngest child consume any yogurt?<br>Any thin porridge?<br>Any other liquids? |
| Did your youngest child drink anything from a bottle with a nipple yesterday during the day or night? |
| Do you remember how old your youngest child was when you started feeding them liquids for the first time? |
| Can you tell me how old your youngest child was in months, when you started feeding them liquids for the first time? |
| At what age should you start to feed your child liquids for the first time? (in months) |
| Please describe everything that your youngest child ate yesterday during the day or night, whether at home or outside the home. Did s/he have any…<br>Food made from grains, such as bread, rice, noodles, porridge<br>Pumpkin, carrots, squash, or sweet potatoes that are yellow or orange inside<br>White potatoes, white yams, manioc, cassava, or any other foods made from roots<br>Any dark green leafy vegetables<br>Any other vegetables?<br>Ripe mangoes, ripe papayas, or other local vitamin A-rich fruits<br>Any other fruits?<br>Liver, kidney, heart, or other organs from domesticated animals such as cow, goat, camel, chicken or duck<br>Any meat from domesticated animals such as beef, camel, lamb, goat, camel, chicken, or duck<br>Liver, kidney, heart, or other organs from wild animals, such as birds, sagaaro, bakayle, wild fowl, wild goat<br>Any flesh from wild animals, such as birds, sagaaro, bakayle, wild fowl, wild goat<br>Eggs<br>Fresh or dried fish, shellfish, or seafood<br>Any foods made from beans, peas, lentils, peanuts, peanut paste or other legumes<br>Any foods made from nuts and seeds such as pumpkin seeds, cashews, jackfruit<br>Cheese, yogurt, or other milk products<br>Any oil, fats, or butter, or foods made with any of these<br>Any sugary foods such as chocolates, sweets, candies, pastries, cakes, or biscuits |

| |
|---|
| Condiments for flavor, such as chilies, spices, herbs, or fish powder<br>Crubs, snails,<br>Foods made with red palm oil, red palm nut, or red palm nut pulp sauce |
| Did your youngest child eat any solid, semi-solid, or soft foods yesterday during the day or at night? |
| Do you remember how old your youngest child was when you started feeding them solid food for the first time? |
| Can you tell me how old your youngest child was in months, when started feeding them solid food for the first time? |
| At what age should you start to feed your child solids for the first time? (in months) |
| MDD-C Scoring<br>Early Initiation<br>Liquids Score<br>Solids Score<br>Total Solid Liquids<br>Exclusive Breastfeeding<br>Group 1: Breastmilk<br>Group 2: Grains, Tubers<br>Group 3: Legumes, Nuts<br>Group 4: Dairy<br>Group 5: Meat<br>Group 6: Eggs<br>Group 7: Vitamin A Rich Fruits & Vegetables<br>Group 8: Other Fruits & Vegetables<br>MDD-C: Total Groups<br>MDD-C: 5 Groups |

Thank you for speaking with us today, we greatly appreciate your time. Do you have any questions for me before I go? If so, I can answer them now.

**Table S5. Midline Survey Instrument**

| **Midline Questionnaire** |
|---|
| **Enumerator and Location Details** |
| Start Time |
| End Time |
| Date |
| Enumerator ID |
| Household Member Unique ID (cash recipient) |
| **According to the HH ID, the respondent is from a household headed by {mother's name} in {village name} in {district name} under {region name}. This household has {number of} child(ren) interviewed at baseline. Please make sure this information is accurate.** |
| Unique ID |
| Displacement Status |
| Region |
| District |
| Village |
| Cluster Number |
| Study Arm |
| **Informed Consent** |
| Hello, we are from Save the Children, and we would like to speak with {mother's name}. Are you {mother's name}? |
| Were you the one who responded during the last interview 3 months ago? |
| What is your relationship to the youngest child in this household?<br>If other, please specify |
| What is your name? |
| What is your age? |
| If not respondent at baseline, administer consent:<br>Thank you for the opportunity to speak with you. As I mentioned, we are from Save the Children. We are conducting a survey with randomly selected program households to learn more about the effect of our multi-purpose cash program. The questions will take about **45 minutes** to complete. Your participation is voluntary. If you agree to participate, you can choose to stop at any time or skip any questions you do not want to answer. There will be no compensation for participating in this survey. If you do not agree to participate, it will not affect your status in the program or the services you receive. Any personal information such as names that you share will be kept confidential and only a limited number of Save the Children staff will have access to it. We intend to share the resulting analysis, which will not include personal information, with our donors and partners to help improve future programs. Let me know if you have any questions about this. |
| Do you agree and consent to be interviewed? |
| Enumerator Name (acknowledging that consent has been obtained) |
| Enumerator Signature (acknowledging that consent has been obtained) |
| Thank you for your time. |
| **Respondent Information** |

| |
|---|
| Respondent’s Telephone Number |
| Respondent’s Alternative Telephone Number |
| Are you currently pregnant? |
| Since we last met you, did you have a baby? |
| **Newborn Details** |
| Name of new baby |
| Who assisted with the delivery of {new baby}?<br>If other, please specify |
| Where did you give birth to {new baby}?<br>If other, please specify |
| Was {new baby} deliver by caesarean (did they cut your belly open to take the baby out? |
| When {new baby} was born, was {new baby} very large, larger than average, average, smaller than average, or very small? |
| **Household Decision Making** |
| Who usually decides how the household income/earnings (all sources of income) will be used: you, your husband, or you and your husband jointly?<br>If other, please specify |
| Who usually makes decisions about healthcare for yourself: you, your husband, or you and your husband jointly?<br>If other, please specify |
| Who usually makes decisions about making major household purchases?<br>If other, please specify |
| In the last 3 months, has any member of your household received such as the government, international organizations, religious bodies in the form of...?<br>None<br>Cash<br>Food, including school feeding<br>Other in-kind, such as animals<br>Micronutrient supplementation (vitamin A, iron, etc)<br>Specialized Nutritious Food (SNF)<br>WASH (hygiene, water, etc)<br>Scholarship<br>Other (please specify) |
| How much did you receive (either total value in USD or kg or other unit)? |
| How often did you receive it? (weekly, monthly, etc) |
| **Basic Causes – Household Assets** |
| Does your household own any of the following livestock?<br>How many camels does your household currently own?<br>How many cattle does your household currently own?<br>How many goats does your household currently own?<br>How many donkeys does your household currently own?<br>How many horses does your household currently own?<br>How many poultry does your household currently own? |

| |
|---|
| Does any member of this household own any agricultural land?<br>How many hectares of agricultural land do members of this household own? |
| Does your household have any of the following? |
| Does any member of this household own:<br>Does any member of this household have a bank account?<br>Does any member of this household use a mobile phone to make financial transactions such as sending or receiving money, paying bills, purchasing goods or services, or receiving wages? |
| How many rooms in this household are used for sleeping? |
| What is the main material of the floor of the house?<br>If other, please specify. |
| What is the main material of the roof of the house?<br>If other, please specify. |
| What is the main material of the exterior walls of the house?<br>If other, please specify. |
| Where do you go to when you have to ease yourself (defecate)?<br>If other, please specify. |
| Where do you dispose of the youngest child's feces?<br>If other, please specify. |
| If you were to give birth to another child, how soon after birth should you begin breastfeeding your child? |
| If you were to give birth to another child, what will you feed the child for the first 6 months of life?<br>If other, please specify. |
| **Household Composition, Morbidity, and Migrations** |
| How many children under 5 years of age are currently living in this household? |
| Enumerator only: record the members of the household that are present during the interview (such as the mother, head of household, children, other family members, etc.) |
| **Child Anthropometrics – Complete for each CU5 in Household** |
| How many children under 5 years of age are currently living in this household? |
| Name of the child |
| Sex of the child |
| What is your relationship to this child? |
| Age of the child (in months) |
| How did you confirm the age?<br>Seasonal Calendar<br>Health card<br>Birth certificate |
| Child's Weight to the nearest 0.1kg |
| Child's Height to the nearest 0.1cm |
| Child's MUAC to the nearest 0.1cm |
| Is oedema present in the child? |
| Mother MUAC |
| Now I am going to take your MUAC measurement from your arm. |

| |
|---|
| Enumerator only: record the members of the household that are present during the interview (such as the mother, head of household, children, other family members, etc) |
| **Household Expenditures** |
| For the next set of questions, I am going to ask you to estimate how much your household spent **in USD** on different household needs over the past month (30 days). |
| How much would you estimate you spent in total on **all your food** over the last 7 days? |
| How much would you estimate you spent in total on **nutritious food** over the last 7 days? |
| In the last month (30 days) how much did your household spend on hygiene products (soap, toothbrush, toothpaste, toilet paper, etc.)? |
| In the last month (30 days) how much did your household spend on transportation (fuel, public transportation, taxi)? |
| In the last month (30 days) how much did your household spend on fuel (wood, paraffin, etc.) for cooking and other needs? |
| In the last month (30 days) how much did your household spend on water? |
| In the last month (30 days) how much did your household spend on lighting or electricity? |
| In the last month (30 days) how much did your household spend on phone credit, internet, or other communication? |
| In the last month (30 days) how much did your household spend on rent or shelter? |
| In the last month (30 days) how much did your household spend on medical costs like doctor's visits and medicine? |
| In the last month (30 days) how much did your child and maternal healthcare like ante- or post-natal care, child vaccinations, etc |
| In the last month (30 days) how much did your household spend on clothing and shoes? |
| In the last month (30 days) how much did your household spend on school fees, school supplies, books, school uniforms, and other education expenses? |
| In the last month (30 days) how much did your household spend on agricultural inputs or livestock and livestock inputs? |
| In the last month (30 days) how much did your household spend on social expenses (gifts, wedding, funeral, etc)? |
| In the last month (30 days) how much did your household spend on debt repayment? |
| In the last month (30 days) how much did your household save? |
| Is it accurate to say your household spent <total> in USD in the past month (30 days)? |
| **Food Consumption Score** |
| I would like to ask you about all the different foods that your household members have eaten in the last 7 days. Could you please tell me how many days in the past week your household has eaten the following foods? |
| In the past week, approximately how many days has your household eaten cereals and tubers such as maize, maize porridge, rice, sorghum, millet, pasta, bread and other cereals, cassava/yucca, potato, or sweet potato? |
| Yes/No? |
| Number of Days |
| In the past week, approximately how many days has your household eaten legumes and nuts such as beans, peas, groundnuts, or cashew nuts? |

| |
|---|
| Yes/No? |
| Number of Days |
| In the past week, approximately how many days has your household eaten vegetables or leaves? |
| Yes/No? |
| Number of Days |
| In the past week, approximately how many days has your household eaten fresh fruits? |
| Yes/No? |
| Number of Days |
| In the past week, approximately how many days has your household eaten eggs or meat such as beef, goat, poultry, camel, or fish? |
| Yes/No? |
| Number of Days |
| In the past week, approximately how many days has your household eaten dairy products such as milk, yogurt, or cheese? |
| Yes/No? |
| Number of Days |
| In the past week, approximately how many days has your household eaten sugar and sugary products such as honey, jam, candy, or pastries. |
| Yes/No? |
| Number of Days |
| In the past week, approximately how many days has your household eaten oils, fats or butter? |
| Yes/No? |
| Number of Days |
| In the past week, approximately how many days has your household eaten condiments such as spices, tea, coffee, salt, fish powder, or small amounts of milk for tea? |
| Yes/No? |
| Number of Days |
| Total FCS Score |
| Category FCS |
| IPC Phase FCS |
| FCS: High Oil |
| IPC Phase FCS: High Oil |
| **Reduced Coping Strategies Index (rCSI)** |
| In the past 7 days, if there have been times when you did not have enough food or money to buy food, how many days has your household had to: |
| a. Rely on less preferred and less expensive foods? |
| Yes/No? |
| Number of Days |
| b. Borrow food, or rely on help from a friend or relative? |
| Yes/No? |
| Number of Days |
| c. Limit portion size at mealtimes? |

| |
|---|
| Yes/No? |
| Number of Days |
| d. Restrict consumption by adults in order for small children to eat? |
| Yes/No? |
| Number of Days |
| e. Reduce number of meals eaten in a day? |
| Yes/No? |
| Number of Days |
| d. Restrict consumption by adults in order for small children to eat? |
| **Household Hunger Scale (HHS)** |
| In the past 30 days, was there ever no food to eat of any kind in your house because of lack of resources to get food?<br>How often did this happen in the past 30 days? |
| In the past 30 days, did you or any household member go to sleep at night hungry because there was not enough food?<br>How often did this happen in the past 30 days? |
| In the past 30 days, did you or any household member go a whole day and night without eating anything at all because there was not enough food?<br>How often did this happen in the past 30 days? |
| HHS Score Total |
| HHS Category |
| HHS IPC Phase |
| **Underlying Causes – Inadequate Care & Feeding Practices** |
| **Child Vaccination History** |
| Thinking about your youngest child, your youngest child, did s/he ever receive any vaccinations, including vaccinations received in campaigns or immunization days or child health days? |
| Has your youngest child ever received any of the following vaccinations?<br>BCG against TB (injection in the arm/shoulder that usually causes a scar)<br>Oral polio vaccine (about 2 drops in the mouth)<br>Pentavalent (injection in the thigh sometimes at the same time as polio drops)<br>Measles vaccination (injection in the arm) |
| How many times did your youngest child receive the pentavalent vaccine? |
| How many times did your youngest child receive the measles vaccine? |
| **Child Health & Nutrition** |
| Has your youngest child had diarrhea, fever, or cough in the last 2 weeks? |
| Did you seek treatment for the illness from any source? |
| **Maternal and Child Wasting** |
| In the last 3 months, did you or your youngest child have your and their height, weight or arm circumference measured in the facility, in the community or in your home? |
| In the last 3 months, were you and your youngest child sent for treatment after the height, weight or arm circumference were measured? |
| If yes, did you or your youngest child receive the treatment? |

| |
|---|
| If yes, what did they receive during the treatment? For example, CSB++ (porridge), Plumpy Nut or medication or anything else?<br>CSB++ (porridge)<br>Plumpy Nut<br>Other, please specify |
| Does the PLW in the HH who have been screened malnutrition have received NFI kits under BHA program? |
| Since the Save Programming has started, have you received any IYCF promotional messages from Save the Children? |
| Did your HH received NFI kits under BHA program? |
| Did you receive the full kit content? |
| If yes, what is your entitlement (quantity in pieces)? |
| If other, please specify. |
| **Underlying causes - Unhealthy HH environment** |
| Now I'm going to ask you about your environment |
| What is the main source of drinking water for members of your household?<br>If other, specify. |
| What do you do to the water before consumption/drinking? (select all that apply)<br>If other, specify. |
| What kind of toilet facility do members of your household usually use?<br>If other, specify. |
| Does the household have access to a handwashing place that has soap and water? |
| What type of handwashing station is it? |
| Is water present for handwashing? |
| Is cleansing agent available for handwashing? |
| When do wash your hands (after what type of daily activities)?<br>Before preparing food<br>Before eating<br>Before feeding children<br>After handling a child's stool/diaper<br>After using latrine/toilet<br>Other (specify) |
| When do you think are the three (3) most important times to wash hands?<br>Before preparing food<br>Before eating<br>Before feeding children<br>After handling a child's stool/diaper<br>After using latrine/toilet<br>Other (specify) |
| SBCC (All study arms) |
| Where should you go to when you have to ease yourself (defecate)?<br>If other, please specify. |
| Where should your child go to when your youngest child wants to ease him/herself (defecate)? |

| |
|---|
| If other, please specify. |
| What should you do to your water before consumption/drinking ? (select all options that apply)<br>If other, please specify. |
| How soon after birth should you begin breastfeeding your child? |
| For the first 6 months of life, tell me all the things a baby should consume |
| SBCC (For Arm 2 – SBCC + Cash) |
| Are there mother to mother support groups in this area? |
| Have you attended any of the mother-to-mother support group sessions? |
| If yes, to what extent was this information helpful in changing the way you will breastfeed? |
| Would you share this information with your neighbors?<br>If no, tell me why. |
| **Immediate Causes - Inadequate Dietary Intake** |
| **Minimum Dietary Diversity for Children (MDD-C)** |
| Has your youngest child ever been breastfed? |
| How long after birth did you put your youngest child to the breast for the first time?<br>Hours?<br>Days? |
| Was your youngest child breastfed yesterday during the day or at night? This includes any breast milk the child drank from a bottle, cup, or spoon as well as directly from the breast.<br>If yes, how many times did you breastfeed your child? |
| Now I would like to ask you about some medicines and vitamins that are sometimes given to infants.<br>Was your youngest child given any vitamin drops or other medicines as drops yesterday during the day or at night? |
| Was your youngest child given oral rehydration solution yesterday during the day or at night? |
| Next, I would like to ask you about some liquids that your youngest child may have had yesterday during the day or at night. Did your youngest child have any of the following…<br>Plain water?<br>Infant formula?<br>How many times yesterday during the day or at night did your youngest child consume any formula?<br>Any milk such as tinned, powdered, or fresh animal milk?<br>How many times yesterday during the day or at night did your youngest child consume any of these types of milk?<br>Any juice or juice drinks?<br>Clear broth?<br>Yogurt?<br>How many times yesterday during the day or at night did your youngest child consume any yogurt?<br>Any thin porridge?<br>Any other liquids? |
| Did your youngest child drink anything from a bottle with a nipple yesterday during the day or night? |

| |
|---|
| Do you remember how old your youngest child was when you started feeding them liquids for the first time? |
| Can you tell me how old your youngest child was in months, when you started feeding them liquids for the first time? |
| At what age should you start to feed your child liquids for the first time? (in months) |
| Please describe everything that your youngest child ate yesterday during the day or night, whether at home or outside the home. Did s/he have any…<br>Food made from grains, such as bread, rice, noodles, porridge<br>Pumpkin, carrots, squash, or sweet potatoes that are yellow or orange inside<br>White potatoes, white yams, manioc, cassava, or any other foods made from roots<br>Any dark green leafy vegetables<br>Any other vegetables?<br>Ripe mangoes, ripe papayas, or other local vitamin A-rich fruits<br>Any other fruits?<br>Liver, kidney, heart, or other organs from domesticated animals such as cow, goat, camel, chicken or duck<br>Any meat from domesticated animals such as beef, camel, lamb, goat, camel, chicken, or duck<br>Liver, kidney, heart, or other organs from wild animals, such as birds, sagaaro, bakayle, wild fowl, wild goat<br>Any flesh from wild animals, such as birds, sagaaro, bakayle, wild fowl, wild goat<br>Eggs<br>Fresh or dried fish, shellfish, or seafood<br>Any foods made from beans, peas, lentils, peanuts, peanut paste or other legumes<br>Any foods made from nuts and seeds such as pumpkin seeds, cashews, jackfruit<br>Cheese, yogurt, or other milk products<br>Any oil, fats, or butter, or foods made with any of these<br>Any sugary foods such as chocolates, sweets, candies, pastries, cakes, or biscuits<br>Condiments for flavor, such as chilies, spices, herbs, or fish powder<br>Foods made with red palm oil, red palm nut, or red palm nut pulp sauce |
| Did your youngest child eat any solid, semi-solid, or soft foods yesterday during the day or at night? |
| Do you remember how old your youngest child was when you started feeding them solid food for the first time? |
| Can you tell me how old your youngest child was in months, when started feeding them solid food for the first time? |
| At what age should you start to feed your child solids for the first time? (in months) |
| MDD-C Scoring<br>Early Initiation<br>Liquids Score<br>Solids Score<br>Total Solid Liquids<br>Exclusive Breastfeeding<br>Group 1: Breastmilk<br>Group 2: Grains, Tubers |

| |
|---|
| Group 3: Legumes, Nuts<br>Group 4: Dairy<br>Group 5: Meat<br>Group 6: Eggs<br>Group 7: Vitamin A Rich Fruits & Vegetables<br>Group 8: Other Fruits & Vegetables<br>MDD-C: Total Groups<br>MDD-C: 5 Groups |
| **Thank you for your time today. Do you have any questions for me before I go? If so, I can answer them now.** |

**Table S6. Endline Survey Instrument**

| # Endline Questionnaire |
|---|
| ***Questions in red are additions to the Endline instrument that were not used at Baseline or Midline*** |
| **Enumerator and Location Details** |
| Start Time |
| End Time |
| Date |
| Enumerator ID |
| Household Member Unique ID (cash recipient) |
| **According to the HH ID, the respondent is from a household headed by {mother's name} in {village name} in {district name} under {region name}. This household has {number of} child(ren) interviewed at baseline. Please make sure this information is accurate.** |
| Unique ID |
| Displacement Status |
| Region |
| District |
| Village |
| Cluster Number |
| Study Arm |
| **Informed Consent** |
| Hello, we are from Save the Children, and we would like to speak with {mother's name}. Are you {mother's name}? |
| Were you the one who responded during the last interview 3 months ago? |
| What is your relationship to the youngest child in this household?<br>If other, please specify |
| What is your name? |
| What is your age? |
| If not respondent at baseline, administer consent:<br>Thank you for the opportunity to speak with you. As I mentioned, we are from Save the Children. We are conducting a survey with randomly selected program households to learn more about the effect of our multi-purpose cash program. The questions will take about **45 minutes** to complete. Your participation is voluntary. If you agree to participate, you can choose to stop at any time or skip any questions you do not want to answer. There will be no compensation for participating in this survey. If you do not agree to participate, it will not affect your status in the program or the services you receive. Any personal information such as names that you share will be kept confidential and only a limited number of Save the Children staff will have access to it. We intend to share the resulting analysis, which will not include personal information, with our donors and partners to help improve future programs. Let me know if you have any questions about this. |
| Do you agree and consent to be interviewed? |
| Enumerator Name (acknowledging that consent has been obtained) |
| Enumerator Signature (acknowledging that consent has been obtained) |

| |
|---|
| Thank you for your time. |
| **Respondent Information** |
| Respondent's Telephone Number |
| Respondent's Alternative Telephone Number |
| What is your current education level? |
| Who is the current head of household? |
| Are you currently pregnant? |
| Since we last met you, did you have a baby? |
| **Newborn Details** |
| Name of new baby |
| Did you attend antenatal care (ANC) while pregnant with this new baby?<br>How many times did you attend antenatal care (ANC)? |
| Did you receive Vitamin A supplementation while pregnant with this new baby? |
| Did you as the mother have any of these illnesses while pregnant with the new baby? |
| Who assisted with the delivery of {new baby}?<br>If other, please specify |
| Where did you give birth to {new baby}?<br>If other, please specify |
| Was {new baby} deliver by caesarean (did they cut your belly open to take the baby out? |
| When {new baby} was born, was {new baby} very large, larger than average, average, smaller than average, or very small? |
| **Current Displacement** |
| Has your household been displaced in the last 3 months?<br>If yes, what is the reason for current displacement? |
| If yes, compared to "baseline" (7 months ago), did the recent displacement in the last 3 months affect your HH access to the following?<br>Money<br>Market<br>Jobs/Livelihood<br>Food<br>Education<br>Healthcare |
| **Household Decision Making** |
| Who usually decides how the household income/earnings (all sources of income) will be used: you, your husband, or you and your husband jointly?<br>If other, please specify |
| Who usually makes decisions about healthcare for yourself: you, your husband, or you and your husband jointly?<br>If other, please specify |
| Who usually makes decisions about making major household purchases?<br>If other, please specify |
| In the last 3 months, has any member of your household received such as the government, international organizations, religious bodies in the form of...?<br>None |

| |
|---|
| Cash<br>Food, including school feeding<br>Other in-kind, such as animals<br>Micronutrient supplementation (vitamin A, iron, etc)<br>Specialized Nutritious Food (SNF)<br>WASH (hygiene, water, etc)<br>Scholarship<br>Other (please specify) |
| How much did you receive (either total value in USD or kg or other unit)? |
| How often did you receive it? (weekly, monthly, etc) |
| **Basic Causes – Household Assets** |
| Does your household own any of the following livestock?<br>How many camels does your household currently own?<br>How many cattle does your household currently own?<br>How many goats does your household currently own?<br>How many donkeys does your household currently own?<br>How many horses does your household currently own?<br>How many poultry does your household currently own? |
| Does any member of this household own any agricultural land?<br>How many hectares of agricultural land do members of this household own? |
| Does your household have any of the following? |
| Does any member of this household own:<br>Does any member of this household have a bank account?<br>Does any member of this household use a mobile phone to make financial transactions such as sending or receiving money, paying bills, purchasing goods or services, or receiving wages? |
| How many rooms in this household are used for sleeping? |
| What is the main material of the floor of the house?<br>If other, please specify. |
| What is the main material of the roof of the house?<br>If other, please specify. |
| What is the main material of the exterior walls of the house?<br>If other, please specify. |
| Where do you go to when you have to ease yourself (defecate)?<br>If other, please specify. |
| Where do you dispose of the youngest child's feces?<br>If other, please specify. |
| If you were to give birth to another child, how soon after birth should you begin breastfeeding your child? |
| If you were to give birth to another child, what will you feed the child for the first 6 months of life?<br>If other, please specify. |
| **Household Composition, Morbidity, and Migrations** |
| How many children under 5 years of age are currently living in this household? |

| |
|---|
| Enumerator only: record the members of the household that are present during the interview (such as the mother, head of household, children, other family members, etc.) |
| **Child Anthropometrics – Complete for each CU5 in Household** |
| How many children under 5 years of age are currently living in this household? |
| Name of the child |
| Sex of the child |
| What is your relationship to this child? |
| Age of the child (in months) |
| How did you confirm the age?<br>Seasonal Calendar<br>Health card<br>Birth certificate |
| Child's Weight to the nearest 0.1kg |
| Child's Height to the nearest 0.1cm |
| Child's MUAC to the nearest 0.1cm |
| Is oedema present in the child? |
| Mother MUAC |
| Now I am going to take your MUAC measurement from your arm. |
| Enumerator only: record the members of the household that are present during the interview (such as the mother, head of household, children, other family members, etc.) |
| **Household Expenditures** |
| For the next set of questions, I am going to ask you to estimate how much your household spent **in USD** on different household needs over the past month (30 days). |
| How much would you estimate you spent in total on **all your food** over the last 7 days? |
| How much would you estimate you spent in total on **nutritious food** over the last 7 days? |
| In the last month (30 days) how much did your household spend on hygiene products (soap, toothbrush, toothpaste, toilet paper, etc.)? |
| In the last month (30 days) how much did your household spend on transportation (fuel, public transportation, taxi)? |
| In the last month (30 days) how much did your household spend on fuel (wood, paraffin, etc.) for cooking and other needs? |
| In the last month (30 days) how much did your household spend on water? |
| In the last month (30 days) how much did your household spend on lighting or electricity? |
| In the last month (30 days) how much did your household spend on phone credit, internet, or other communication? |
| In the last month (30 days) how much did your household spend on rent or shelter? |
| In the last month (30 days) how much did your household spend on medical costs like doctor's visits and medicine? |
| In the last month (30 days) how much did your child and maternal healthcare like ante- or post-natal care, child vaccinations, etc |
| In the last month (30 days) how much did your household spend on clothing and shoes? |
| In the last month (30 days) how much did your household spend on school fees, school supplies, books, school uniforms, and other education expenses? |

| |
|---|
| In the last month (30 days) how much did your household spend on agricultural inputs or livestock and livestock inputs? |
| In the last month (30 days) how much did your household spend on social expenses (gifts, wedding, funeral, etc)? |
| In the last month (30 days) how much did your household spend on debt repayment? |
| In the last month (30 days) how much did your household save? |
| Is it accurate to say your household spent <total> in USD in the past month (30 days)? |
| **Food Consumption Score** |
| I would like to ask you about all the different foods that your household members have eaten in the last 7 days. Could you please tell me how many days in the past week your household has eaten the following foods? |
| In the past week, approximately how many days has your household eaten cereals and tubers such as maize, maize porridge, rice, sorghum, millet, pasta, bread and other cereals, cassava/yucca, potato, or sweet potato? |
| Yes/No? |
| Number of Days |
| In the past week, approximately how many days has your household eaten legumes and nuts such as beans, peas, groundnuts, or cashew nuts? |
| Yes/No? |
| Number of Days |
| In the past week, approximately how many days has your household eaten vegetables or leaves? |
| Yes/No? |
| Number of Days |
| In the past week, approximately how many days has your household eaten fresh fruits? |
| Yes/No? |
| Number of Days |
| In the past week, approximately how many days has your household eaten eggs or meat such as beef, goat, poultry, camel, or fish? |
| Yes/No? |
| Number of Days |
| In the past week, approximately how many days has your household eaten dairy products such as milk, yogurt, or cheese? |
| Yes/No? |
| Number of Days |
| In the past week, approximately how many days has your household eaten sugar and sugary products such as honey, jam, candy, or pastries. |
| Yes/No? |
| Number of Days |
| In the past week, approximately how many days has your household eaten oils, fats or butter? |
| Yes/No? |
| Number of Days |
| In the past week, approximately how many days has your household eaten condiments such as spices, tea, coffee, salt, fish powder, or small amounts of milk for tea? |

| |
|---|
| Yes/No? |
| Number of Days |
| Total FCS Score |
| Category FCS |
| IPC Phase FCS |
| FCS: High Oil |
| IPC Phase FCS: High Oil |
| **Reduced Coping Strategies Index (rCSI)** |
| In the past 7 days, if there have been times when you did not have enough food or money to buy food, how many days has your household had to: |
| a. Rely on less preferred and less expensive foods? |
| Yes/No? |
| Number of Days |
| b. Borrow food, or rely on help from a friend or relative? |
| Yes/No? |
| Number of Days |
| c. Limit portion size at mealtimes? |
| Yes/No? |
| Number of Days |
| d. Restrict consumption by adults in order for small children to eat? |
| Yes/No? |
| Number of Days |
| e. Reduce number of meals eaten in a day? |
| Yes/No? |
| Number of Days |
| d. Restrict consumption by adults in order for small children to eat? |
| **Household Hunger Scale (HHS)** |
| In the past 30 days, was there ever no food to eat of any kind in your house because of lack of resources to get food?<br>How often did this happen in the past 30 days? |
| In the past 30 days, did you or any household member go to sleep at night hungry because there was not enough food?<br>How often did this happen in the past 30 days? |
| In the past 30 days, did you or any household member go a whole day and night without eating anything at all because there was not enough food?<br>How often did this happen in the past 30 days? |
| HHS Score Total |
| HHS Category |
| HHS IPC Phase |
| **Underlying Causes – Inadequate Care & Feeding Practices** |
| **Child Vaccination History** |
| Thinking about your youngest child, your youngest child, did s/he ever receive any vaccinations, including vaccinations received in campaigns or immunization days or child health days? |

| |
|---|
| Has your youngest child ever received any of the following vaccinations?<br>BCG against TB (injection in the arm/shoulder that usually causes a scar)<br>Oral polio vaccine (about 2 drops in the mouth)<br>Pentavalent (injection in the thigh sometimes at the same time as polio drops)<br>Measles vaccination (injection in the arm) |
| How many times did your youngest child receive the pentavalent vaccine? |
| How many times did your youngest child receive the measles vaccine? |
| **Child Health & Nutrition** |
| Has your youngest child had diarrhea, fever, or cough in the last 2 weeks? |
| Did you seek treatment for the illness from any source? |
| **Maternal and Child Wasting** |
| In the last 3 months, did you or your youngest child have your and their height, weight or arm circumference measured in the facility, in the community or in your home? |
| In the last 3 months, were you and your youngest child sent for treatment after the height, weight or arm circumference were measured? |
| If yes, did you or your youngest child receive the treatment? |
| If yes, what did they receive during the treatment? For example, CSB++ (porridge), Plumpy Nut or medication or anything else?<br>CSB++ (porridge)<br>Plumpy Nut<br>Other, please specify |
| Does the PLW in the HH who have been screened malnutrition have received NFI kits under BHA program? |
| Since the Save Programming has started, have you received any IYCF promotional messages from Save the Children? |
| Did your HH received NFI kits under BHA program |
| Did you received the full kit content? |
| If yes, what is your entitlement (quantity in pieces)? |
| If other, please specify. |
| **Underlying causes - Unhealthy HH environment** |
| Now I'm going to ask you about your environment |
| What is the main source of drinking water for members of your household?<br>If other, specify. |
| What do you do to the water before consumption/drinking? (select all that apply)<br>If other, specify. |
| What kind of toilet facility do members of your household usually use?<br>If other, specify. |
| Does the household have access to a handwashing place that has soap and water? |
| What type of handwashing station is it? |
| Is water present for handwashing? |
| Is cleansing agent available for handwashing? |
| When do wash your hands (after what type of daily activities)?<br>Before preparing food |

| |
|---|
| Before eating<br>Before feeding children<br>After handling a child's stool/diaper<br>After using latrine/toilet<br>Other (specify) |
| When do you think are the three (3) most important times to wash hands?<br>Before preparing food<br>Before eating<br>Before feeding children<br>After handling a child's stool/diaper<br>After using latrine/toilet<br>Other (specify) |
| SBCC (All study arms) |
| Where should you go to when you have to ease yourself (defecate)?<br>If other, please specify. |
| Where should your child go to when your youngest child wants to ease him/herself (defecate)?<br>If other, please specify. |
| What should you do to your water before consumption/drinking ? (select all options that apply)<br>If other, please specify. |
| How soon after birth should you begin breastfeeding your child? |
| For the first 6 months of life, tell me all the things a baby should consume |
| SBCC (For Arm 2 – SBCC + Cash) |
| Are there mother to mother support groups in this area? |
| Have you attended any of the mother-to-mother support group sessions?<br>If yes, to what extent was this information helpful in changing the way you will breastfeed? |
| If yes, to what extent was this information helpful in changing the way you will breastfeed? |
| Would you share this information with your neighbors?<br>If no, tell me why. |
| **Immediate Causes - Inadequate Dietary Intake** |
| **Minimum Dietary Diversity for Children (MDD-C)** |
| Has your youngest child ever been breastfed? |
| How long after birth did you put your youngest child to the breast for the first time?<br>Hours?<br>Days? |
| Was your youngest child breastfed yesterday during the day or at night? This includes any breast milk the child drank from a bottle, cup, or spoon as well as directly from the breast.<br>If yes, how many times did you breastfeed your child? |
| Now I would like to ask you about some medicines and vitamins that are sometimes given to infants.<br>Was your youngest child given any vitamin drops or other medicines as drops yesterday during the day or at night? |
| Was your youngest child given oral rehydration solution yesterday during the day or at night? |

Next, I would like to ask you about some liquids that your youngest child may have had yesterday during the day or at night. Did your youngest child have any of the following…

- Plain water?
- Infant formula?
  - How many times yesterday during the day or at night did your youngest child consume any formula?
- Any milk such as tinned, powdered, or fresh animal milk?
  - How many times yesterday during the day or at night did your youngest child consume any of these types of milk?
- Any juice or juice drinks?
- Clear broth?
- Yogurt?
  - How many times yesterday during the day or at night did your youngest child consume any yogurt?
- Any thin porridge?
- Any other liquids?

Did your youngest child drink anything from a bottle with a nipple yesterday during the day or night?

Do you remember how old your youngest child was when you started feeding them liquids for the first time?

Can you tell me how old your youngest child was in months, when you started feeding them liquids for the first time?

At what age should you start to feed your child liquids for the first time? (in months)

Please describe everything that your youngest child ate yesterday during the day or night, whether at home or outside the home. Did s/he have any…

- Food made from grains, such as bread, rice, noodles, porridge
- Pumpkin, carrots, squash, or sweet potatoes that are yellow or orange inside
- White potatoes, white yams, manioc, cassava, or any other foods made from roots
- Any dark green leafy vegetables
- Any other vegetables?
- Ripe mangoes, ripe papayas, or other local vitamin A-rich fruits
- Any other fruits?
- Liver, kidney, heart, or other organs from domesticated animals such as cow, goat, camel, chicken or duck
- Any meat from domesticated animals such as beef, camel, lamb, goat, camel, chicken, or duck
- Liver, kidney, heart, or other organs from wild animals, such as birds, sagaaro, bakayle, wild fowl, wild goat
- Any flesh from wild animals, such as birds, sagaaro, bakayle, wild fowl, wild goat
- Eggs
- Fresh or dried fish, shellfish, or seafood
- Any foods made from beans, peas, lentils, peanuts, peanut paste or other legumes
- Any foods made from nuts and seeds such as pumpkin seeds, cashews, jackfruit
- Cheese, yogurt, or other milk products
- Any oil, fats, or butter, or foods made with any of these
- Any sugary foods such as chocolates, sweets, candies, pastries, cakes, or biscuits

| |
|---|
| Condiments for flavor, such as chilies, spices, herbs, or fish powder<br>Foods made with red palm oil, red palm nut, or red palm nut pulp sauce |
| Did your youngest child eat any solid, semi-solid, or soft foods yesterday during the day or at night? |
| Do you remember how old your youngest child was when you started feeding them solid food for the first time? |
| Can you tell me how old your youngest child was in months, when started feeding them solid food for the first time? |
| At what age should you start to feed your child solids for the first time? (in months) |
| MDD-C Scoring<br>Early Initiation<br>Liquids Score<br>Solids Score<br>Total Solid Liquids<br>Exclusive Breastfeeding<br>Group 1: Breastmilk<br>Group 2: Grains, Tubers<br>Group 3: Legumes, Nuts<br>Group 4: Dairy<br>Group 5: Meat<br>Group 6: Eggs<br>Group 7: Vitamin A Rich Fruits & Vegetables<br>Group 8: Other Fruits & Vegetables<br>MDD-C: Total Groups<br>MDD-C: 5 Groups |
| **Thank you for your time today. Do you have any questions for me before I go? If so, I can answer them now.** |

**Table S7. Exposures: Household Survey Indicators and Definitions**

| | |
|---|---|
| **Enabling Causes** | |
| *Context, Capital, and Resources* | |
| Demographic Information and Household Characteristics | Participants were surveyed on a variety of household and demographic characteristics including:<br>- Sex of the Head of Household<br>- Displacement due to flooding (endline)<br>- Receipt of Cash Assistance/ Livelihood Support from other programs |
| Maternal Education | Mothers were asked what their highest level of education was:<br>- No formal education<br>- Madrasa<br>- Primary<br>- Secondary |
| Maternal Empowerment and Decision-making | Mothers were asked about their involvement in household decision-making in three areas:<br>- Income<br>- Healthcare<br>- Purchases<br>For each area, the decision-maker was either:<br>- Father<br>- Mother<br>- Joint decision-making |
| Household Assets: Principal Component Analysis | Household assets and wealth were evaluated using the Principal Component Analysis index. The index uses asset indicators rather than income data to visualize long-term economic status.<br><br>The asset index considered ownership of the following assets and housing characteristics:<br>- Livestock ownership<br>- Land ownership (in hectares)<br>- Back account ownership<br>- Household items (electricity, radio, TV, telephone, computer, refrigerator, internet air conditioning)<br>- HH Assets (watch, mobile phone, bicycle, scooter, donkey cart, truck, canoe, tractor, ox plough)<br>- Crowding status<br>- Floor<br>- Roof |

| | |
|---|---|
| | - Walls<br>- Type of toilet<br>- Water Source<br>Household were then categorized into asset index quintiles: poorest, poorer, middle, richer, and richest. |
| Household Expenditures | Participants were surveyed on monthly expenditures (absolute dollar value) and expenditure categories.<br><br>Expenditure categories included: food, water, medical needs, agriculture, hygiene, electricity, maternal/child health needs, social activities, transport, communication, clothes and NFIs, debt repayment, fuel, rent, school fees, and savings. |
| **Underlying Causes** | |
| *Household Food Security* | |
| Household Hunger Scale | The Household Hunger Scale (HHS) measures the scale of households' food deprivation.<br><br>Method: Participants are asked if their HH has experienced any of the following in the past 30 days and how often?<br>- No food to eat of any kind because of lack of resources to get food.<br>- A HH member had to sleep hungry because there was not enough food.<br>- A HH member had to go a whole day and night without eating anything at all because there was not enough food.<br><br>Frequencies are coded as rarely (1-2 times), sometimes (3-10 times), and often (>10 times).<br><br>Scores are summed and categorized as:<br>0-1: Little to no hunger<br>2-3: Moderate hunger<br>4-6: Severe hunger |
| Food Consumption Score | Measures how often household consume food items from the different food groups during a 7-day reference period. |

<table>
<tr><td></td><td>Method: The consumption frequency of each food group is multiplied by its weight.<br><br>
<table>
<tr><th></th><th>Food Group</th><th>Weight</th></tr>
<tr><td>1</td><td>Main staples</td><td>2</td></tr>
<tr><td>2</td><td>Pulses</td><td>3</td></tr>
<tr><td>3</td><td>Vegetables</td><td>1</td></tr>
<tr><td>4</td><td>Fruit</td><td>1</td></tr>
<tr><td>5</td><td>Meat/Fish</td><td>4</td></tr>
<tr><td>6</td><td>Dairy</td><td>4</td></tr>
<tr><td>7</td><td>Sugar</td><td>0.5</td></tr>
<tr><td>8</td><td>Oils</td><td>0.5</td></tr>
</table>
<br>Scores are summed and categorized as:<br>0-28: Poor<br>28-35: Borderline<br>>35: Acceptable</td></tr>
<tr><td>Reduced Coping Strategies Index</td><td>Measures the frequency and severity of HH food consumption behaviors due to food shortage in the 7 days prior.<br><br>Method: Participants are asked a series of 5 weighted questions and how often they have experienced these in the past 7 days.<br><br>
<table>
<tr><th>Question</th><th>Weight</th></tr>
<tr><td>Relies on less preferred food</td><td>1</td></tr>
<tr><td>Borrows food/or rely on friends &relatives</td><td>2</td></tr>
<tr><td>Limits portion size at mealtimes</td><td>1</td></tr>
<tr><td>Restricts consumption by adults</td><td>3</td></tr>
<tr><td>Reduce the # of meals in a day</td><td>1</td></tr>
</table>
<br>Scores are summed and range from 0-56. Higher scores mean households increasingly use worse coping mechanisms and have increased food insecurity.</td></tr>
<tr><td colspan="2">Unhealthy Household Environment</td></tr>
<tr><td>Household Crowding</td><td>Participants provided a household roster at survey timepoints.<br><br>Household crowding was defined as:<br>Not crowded: Less 5 people in the HH<br>Crowded: 5 or more people in the HH</td></tr>
</table>

| | |
|---|---|
| | Mothers also reported the number of CU5 residing in the household. |
| Water, Sanitation, and Hygiene Practices (WaSH) | Participants were asked questions about their access to:<br>- Handwashing facilities with water and soap<br>- Improved/unimproved sanitation facilities<br>- Improved/unimproved water sources<br><br>Sanitation facilities and water sources were classified as improved or unimproved based on the DHS classification.<br><br>Sanitation:<br>Improved: flushing toilet, pit latrine, composting toilet<br>Unimproved: open defecation in a bush or field, bucket, other<br><br>Water Source:<br>Improved: piped water, public tap/standpipe, protected well, protected spring, rainwater, tanker truck, cart with small tank<br>Unimproved: unprotected well, unprotected spring, surface water, or other |
| *Inadequate Care and Feeding Practices* | |
| Maternal and Child Health | Participants were surveyed on topics related to maternal and child health including place of last birth, whether their last birth was assisted by a skilled-birthing attendant, caesarean birth, and child vaccination history. |
| Malnutrition Screening | Participants were asked if they or their child received malnutrition screening in a health facility, in the community, or at home in the last 3 months, whether they were referred for malnutrition treatment, and whether their child is currently enrolled in wasting treatment. |
| SBCC Indicators | Participants were surveyed on topics related to knowledge and practice of health and ICYF behaviors, including initiation and exclusivity of breastfeeding, initiation of liquid and solid foods, water treatment, and critical handwashing moments. Arm 2 participants |

| | |
|---|---|
| | were also surveyed on awareness of M2M support groups. |
| **Intervention Level Causes** | |
| Other Inventions/Cash Assistance | Participants were asked whether they were currently receiving support from other cash assistance or nutrition support programs. |
| **Immediate Causes** | |
| *Inadequate Dietary Intake* | |
| Minimum Dietary Diversity – Child | Measures how many of these food groups the child consumed the day before survey data collection:<br>- Breastmilk<br>- Grains, roots, tubers<br>- Legumes, nuts<br>- Dairy products<br>- Flesh foods (meat/fish)<br>- Eggs<br>- Vitamin A rich fruits and vegetables<br>- Other fruits and vegetables<br><br>Scores range from 0 (consumed none of these food groups) to 8 (consumed all of these food groups) and dietary diversity is categorized as:<br>- Does not meet dietary diversity: <5 food groups<br>- Meets dietary diversity: 5 or more food groups |
| Maternal Characteristics | Maternal characteristics relevant to child wasting:<br>- Age<br>- Pregnancy status<br>- Maternal wasting, measured by MUAC |
| *Diseases* | |
| Childhood Illness | Participants were asked whether their child experienced diarrhea, cough, or fever, within the past two weeks. |

### Annex S8. Mothers’ Focus Group Discussion Guides

#### FGD Guide Draft Questions- Mothers

**Objective:** The goal of these focus group discussions is to offer additional insight and perspective into the quantitative baseline results, probing into thematic areas of interest to better understand participants’ experiences and perceptions as well as key barriers and facilitators surrounding cash for nutrition and prevention of wasting in the study area.

**Themes:**

Program
Use of Cash
Health-Seeking Behaviors during Pregnancy
Child Sickness + Health-Seeking Behaviors when Sick
Dietary Diversity + Food Security

**Guide:**

***Program***

**1a.** Tell me about the cash plus intervention in your area. How would you describe recipients’ experiences with receiving cash assistance?

- **1b.** How would you describe the program?
- **1c.** What was good and what was not good?
- **1d.** What worked well and what did not work well?
- **1e.** What could improve the program?
- **1f.** When receiving cash, is it possible for households to face any other difficulties and hardships? If so, what are these difficulties?

**2a.** SBCC: Have you received messaging around cash use, food, and nutrition? If so, was this messaging helpful?

- **2b.** Was there enough messaging?
- **2c.** Do you think this messaging helps people receiving cash change their practices?
- **2d.** Among those who attended educational sessions: Did you attend any educational sessions? If so, describe the educational sessions you attended.
- **2e.** What do you think the SBCC participants learned? What do you think they found useful about the sessions?
- **2f.** What did participants enjoy or not enjoy about the sessions?
- **2g.** What could be improved?

***Use of Cash***

**3a.** Now we’d like to discuss more about how people use the cash that was given as part of the BHA program. What were the main purchases the cash was used for?

- **3b.** Probe: For each main purchase listed, probe into why.
- **3c.** What is good about receiving cash? What is bad about receiving cash?
    - **3d.** Could receiving cash cause any conflict or tension within the house, community, etc.?
- **3e.** In this community, who in the household typically decides how the cash was used?

**3f.** Do women have influence over how the cash is used? Do women in the community get to decide how the cash is used?

**3g.** Is the cash assistance enough for families in the community to be able to purchase adequate food for their households?

**3h.** Is the cash amount sufficient to meet household needs?

**3i.** Are recipients able to purchase the types of foods that they normally would consume?

**3j.** Do recipients of cash purchase different things at different times in the year?

**3k.** Now thinking about coping strategies, what coping strategies are generally used to pay off debt?

***Health-Seeking Behaviors during Pregnancy***

**4a.** We're now interested in discussing pregnancy and health practices during pregnancy in the community. How do women in the community take care of themselves during pregnancy?

**4b.** What should women eat when they are pregnant?

**4c.** What diseases are most common among pregnant women?

**4d.** Probe into other beliefs, taboos, etc. around pregnancy: Should pregnant women eat anything in particular?

**4e.** What should pregnant women do to stay healthy and have a healthy baby?

**4f.** Should a woman go to a health center during pregnancy? What about if women are sick while they are pregnant?

**4g.** Where is it common for women in the community to have their baby?

***Child Sickness + Health-Seeking Behaviors when Sick***

**5a.** Can you now talk to me about what a child needs to be healthy?

**5b.** What about when they are sick?

**5c.** What do mothers in the community worry about surrounding the health of their children?

**5d.** Do people in the community face any difficulties in accessing health services? If so, what are these challenges?

**5e.** What are some common beliefs, taboos, practices, etc. surrounding caring for a sick child: Does a child eat anything special or different when they are sick?

**5f.** How should a child be cared for if they are sick?

**5g.** What should be done if a child has diarrhea or a fever?

**5h.** What makes a child get sick?

**5i.** What is meant by the term "wasting"? What causes wasting?

**5j.** What are the community's views of wasting?

**5k.** What should happen if a child is wasted?

***Dietary Diversity + Food Security***

**6a.** What foods are typically consumed by households in the community? Why?

**6b.** What foods are available in the area?

**6c.** Do people in the community face any difficulty in accessing any of these foods?

**6d.** Are there foods that people might want but that they do not have access to? Why is that?

**6e.** Probe into access to markets- are markets easily accessible in the community?

**6f.** Do community members face any challenges or barriers in accessing markets?

**6g.** Why is it important to eat different types of foods?

**7a.** So now thinking about infants under 6 months of age, what are these children being fed?

**7b.** What about children that are older than 6 months of age? What are they commonly being fed?

**7c.** How many times a day do children eat?

**7d.** What challenges do mothers in your community face feeding their children?

**8a.** Talk to me about breastfeeding in your community. Do women commonly breastfeed?

**8b.** Do they have any challenges with breastfeeding and, if so, what are they?

### Annex S9. Men's Focus Group Discussion Guide

#### FGD Guide Draft Questions- Men

**Objective:** The goal of these focus group discussions is to offer additional insight and perspective into the quantitative baseline results, probing into thematic areas of interest to better understand participants' experiences and perceptions as well as key barriers and facilitators surrounding cash for nutrition and prevention of wasting in the study area.

**Themes:**

Program
Use of Cash
Supporting Healthy Pregnancy
Child Sickness + Health-Seeking Behaviors when Sick
Dietary Diversity + Food Security

**Guide:**

***Program***

**1a.** Tell me about the cash plus intervention in your area. How would you describe recipients' experiences with receiving cash assistance?

- **1b.** How would you describe the program?
- **1c.** What was good and what was not good?
- **1d.** What worked well and what did not work well?
- **1e.** What could improve the program?
- **1f.** When receiving cash, is it possible for households to face any other difficulties and hardships? If so, what are these difficulties?

**2a.** Did anyone in your household receive messages related to cash use, food, and nutrition? If yes, can you tell me about what messages they told you they received?

***Use of Cash***

**3a.** Now we'd like to discuss more about how people use the cash that was given as part of the BHA program. Do you know how the cash was used? If so, what were the main purchases the cash was used for?

- **3b.** Probe: For each main purchase listed, probe into why.
- **3c.** What is good about receiving cash? What is bad about receiving cash?
  - **3d.** Could receiving cash cause any conflict or tension within the house, community, etc.?
- **3e.** In this community, who in the household typically decides how the cash was used?
  - **3f.** What are men's roles in deciding how to use the cash?
  - **3g.** Is there agreement in households on how cash should be used?
  - **3h.** Do different family members think the cash should be used differently?
- **3i.** Is the cash assistance enough for families in the community to be able to purchase adequate food for their households?

**3j.** Is the cash amount sufficient to meet household needs?

**3k.** Are recipients able to purchase the types of foods that they normally would consume?

**3l.** Now thinking about coping strategies, what coping strategies are generally used to pay off debt?

***Supporting Healthy Pregnancy***

**4a.** We're now interested in discussing pregnancy and health practices during pregnancy in the community.

**4b.** What types of things do men in the community do to support their partners while they're pregnant?

**4c.** What sort of things are important for a healthy pregnancy?

***Child Sickness + Health-Seeking Behaviors when Sick***

**5a.** Can you now talk to me about what a child needs to be healthy? What about when they are sick?

**5b.** What are men's responsibilities surrounding childcare?

**5c.** In what ways are men's childcare roles different from women's?

**5d.** In this community, if a father was more involved in child care activities than usual, what would other fathers think of this?

**5e.** What makes a child get sick?

**5f.** What is meant by the term "wasting"? What causes wasting?

**5g.** What are the community's views of wasting?

**5h.** What should happen if a child is wasted?

**5i.** What should be done to prevent wasting?

***Dietary Diversity + Food Security***

**6a.** What foods are typically consumed by households in the community? Why?

**6b.** What are men's roles in deciding what foods are consumed in the household?

**6c.** Do men eat any foods specifically that other members of the family do not eat?

**Table S10. Market Monitoring Data Collection Sites**

| Region | Market Monitoring Data Collection Areas |
|---|---|
| *Bay* | Barxada<br>Hanano<br>Ondon<br>Adaada<br>Dugandug<br>Baidoa |
| *Hiran* | Yoobsan/daraawiish<br>Jawil<br>Mahas<br>Gerjiir<br>Beegadid<br>Mataban |

**Table S11. Food Commodities Monitored During Market Monitoring**

| Category | Foods |
|---|---|
| *Sentinel Foods* | Sorghum flour, white rice, pasta, wheat flour, imported rice, vegetable oil |
| *Fruits and Vegetables* | Banana, Swiss chard, groundnuts, pumpkin, spinach, potatoes, lime, onion |
| *Cereals* | Beans, lentils, maize, whaite maize, sorghum, red sorghum, cowpeas, baits (canned) |
| *Protein Sources* | Meat, eggs, cooked eggs, goat liver, canned sardines, canned milk, camel meat, camel milk |

**Table S12. Market Monitoring Questionnaire**

| Market Monitoring Questionnaire |
|---|
| **Enumerator and Location Details** |
| Start Time |
| End Time |
| Date |
| Interviewer Name |
| Goods: What type of goods are you (the interviewer) collecting information on today? Select all that apply.<br>Food<br>WASH NFIs<br>Rent<br>Household goods or other shelter NFIs<br>Childcare Products<br>Other |
| Region |
| District |
| Administrative Level 1: Department/State<br>Bay<br>Hiran |
| Administrative Level 2: Municipality, District, etc.<br>Beledweyne<br>Mahas<br>Mataban<br>Baidoa |
| Administrative Level 3: Community, Town, Village<br>Yoobsan/daraawiish<br>Jawil<br>Mataban Town<br>Bergadid<br>Gerijir<br>Mahas Town<br>Baidoa Town<br>Ondon<br>Dugandug<br>Adada<br>Barxadda<br>Hanano<br>Other |
| Market Name:<br>Intervention market 1 Yoobsan/daraawiish<br>Intervention market 2 Jawil<br>Intervention market 3 Bergadid<br>Intervention market 4 Mataban Town |

Intervention market 5 Gerijir
Intervention market 6 Mahas Town
Intervention market 7 Bergadid
Intervention market 8 Ondon
Intervention market 9 Dugandug
Intervention market 10 Adada
Intervention market 11 Barxadda
Intervention market 12 Hanano
Regional supply market 1 Beledweyne
Regional supply market 2 Baidoa
Central market 1 Beledweyne Market
Central market 2 Baidoa Market
Comparison market 1
Comparison market 2

**Informed Consent**

Thanks for the opportunity to talk to you. We are from Save the Children. We are conducting a survey with randomly selected vendors to learn more about the markets near our multi-purpose cash program. We collect information about prices and weights for some products that are regularly purchased by the participants. We intend to use the information to determine whether our programs are affecting markets and prices in a positive or negative way. The questions will take about 20 minutes to complete, and in some cases we may ask to weigh products with our scales.

Your participation is entirely voluntary. If you agree to participate, you can choose to stop at any time or skip any questions you do not want to answer. There will be no compensation of any kind for participating in this survey.

Any personally identifiable information (such as names and phone numbers) that you share will be kept confidential. A limited number of [name of organization] staff will have access to it so they can contact you remotely if they need to ask follow up questions. We intend to share the resulting analysis, which will not include personally identifiable information, with the donor and partners to help improve programs. Let me know if you have any questions about this.

If you have additional questions after this, you can also contact [enter staff member name and a phone number].

Do you agree to speak with us?

Enumerator Name (acknowledging that consent has been obtained)

Enumerator Signature (acknowledging that consent has been obtained)

Thank you for your time.

**Vendor Information**

Vendor Name

Vendor Actor Type:
Importer
Provincial Trader
District Trader

| |
|---|
| Urban Retailer<br>Village Trader<br>Commercial Farmer<br>Wholesaler<br>Other |
| Price Type<br>Market Prices<br>Wholesale Prices |
| Vendor Contact and Phone Number |
| **Food** |
| For enumerator: Select which unit your scale uses.<br>Grams (g)<br>Kilograms (kg)<br>Ounces (oz)<br>Pounds (lb)<br>Other (please specify) |
| Which of the following food commodities do you have in stock today?<br>Beans<br>Lentils<br>Maize<br>Sorghum<br>Sorghum Flour<br>White Rice<br>Meat<br>Red Sorghum<br>Banana<br>Other Fruits<br>Milk (powdered and animal sources)<br>Porridge<br>Pumpkin Seeds<br>Groundnuts<br>Swiss Chard<br>Eggs (chicken, cooked)<br>Pumpkin<br>Baits (oats)<br>Citrus Fruits<br>Pasta<br>White Sorghum<br>Wheat Flour<br>Whaite Maize<br>Imported Rice<br>Canned Sardines<br>Goat Liver<br>Goat Meat<br>Camel Meat<br>Camel Milk |

Eggs
Spinach
Cowpeas
Potatoes
Lime
Vegetable Oil
Onion

***For each commodity the vendor has in stock, ask the following questions:***

What is the most common unit of sale for {commodity}?
What is the sale price of one unit of {commodity}?

***For enumerator:***

Please weigh the unit of {commodity} with your scale and write the weight shown on the scale.

**For each commodity, the data evaluation team will then calculate:**

Market Price per Unit
Minimum Expenditure Basket: monthly cost per household, based on monthly quantity of {commodity} per household

**WASH NFIs**

Which of the following {item}s do you have in stock today?

Bucket
Jerrycan
Clothes washing basin
Body soap
Laundry soap
Toothbrush
Toothpaste
Sanitary pads
Other wash items as applicable

***For each item the vendor has in stock, ask the following questions:***

What is the sale price of one unit of {item}?

**For each item, the data evaluation team will then calculate:**

Minimum Expenditure Basket: monthly cost per household, based on monthly quantity of units of {item} per household

**Childcare NFIs**

Which of the following {item}s do you have in stock today?

Children's diapers
Anti-itch cream
Other childcare items as applicable

***For each item the vendor has in stock, ask the following questions:***

What is the sale price of one unit of {item}?

**For each item, the data evaluation team will then calculate:**

Minimum Expenditure Basket: monthly cost per household, based on monthly quantity of units of {item} per household

**Shelter and Household NFIs**

| |
|---|
| Which of the following {item}s do you have in stock today?<br>Pots<br>Flat dish (plate, stainless steel)<br>Metal utensils<br>Scoop/ladle (aluminum)<br>Electric Stove<br>Bedsheets<br>Blankets<br>Mattress/sleeping mat<br>Plastic sheets/tarpaulin<br>Rope |
| ***For each item the vendor has in stock, ask the following questions:***<br>What is the sale price of one unit of {item}? |
| **For each item, the data evaluation team will then calculate:**<br>Minimum Expenditure Basket: monthly cost per household, based on monthly quantity of units of {item} per household |
| **Market Supply** |
| **Availability:** Has availability for any of these products changed noticeably compared to the same time last year?<br>For which products?<br>What do you think are the reasons for changing availability? |
| **Stock:** Do you normally have any of the items we discussed in stock, but you're out of stock today? As a reminder, we talked about [ENUMERATOR to list items discussed].<br>Which items?<br>Why are you out of stock?<br>When was the last time you had these commodities in stock? |
| Price Changes: Have you noticed a significant change in price for any of these items within the past month?<br>For which products?<br>Why do you think the price has changed? |
| **Thank you for speaking with us and have a good day.** |

### Annex S13. Research Uptake Strategy

#### Strategic Objectives for Stakeholder Engagement

In order to contribute to the aforementioned high-level **research impact objective,** what we hope to deliver over the next 1-2 years is to adapt SC's cash for nutrition programming and inform the design of cash for relapse prevention interventions in similar contexts. We expect that this short-term impact will contribute to the delivery of the high-level impact objective because we will implement evidence-based interventions to ensure we are providing the most appropriate, effective, and cost-effective services. Additionally, we aim to impact global stakeholders so that other implementing partners programming cash for nutrition will consider new evidence in decision-making and program roll-out.

More specifically, through this research:

3) Global and regional level stakeholders will become aware of the long-term positive impacts of nutrition-sensitive cash programming in humanitarian settings and use our evidence to inform how they fund and design cash plus nutrition interventions to prevent acute malnutrition and relapse. The evidence will also be used to prioritize and standardize cash, nutrition, and SBCC indicators, and influence nutrition SBCC strategies through the Global Nutrition Cluster.
3) Key Somali stakeholders, including the Ministry of Health and National Bureau of Statistics, will use our evidence to inform/adapt policies and programming on cash plus nutrition at the national and subnational level, namely the National Health Plan and Nutrition Policy; and to help mobilize funding of humanitarian-based cash plus nutrition programs to prevent malnutrition.
3) Save the Children will use the evidence to adapt the Resourcing Families for Better Nutrition (RF4BN) or cash plus nutrition programming in humanitarian settings as well as to inform design of cash plus nutrition projects and additional research to prevent relapse among children under five. SC will share key lessons learned and best practices on the design of cash plus nutrition programming with other implementing partners through the Global Nutrition Cluster and CALP Network.

#### Target Stakeholders

After analysis and mapping, target stakeholders were identified from these organizations: Somalia Federal Ministry of Health (MoH), Somalia Federal Ministry of Labour and Social Affairs, Somalia Office of the Prime Minster, UNICEF, WFP, Concern Worldwide, Save the Children International Somalia, Save the Children International, USAID/BHA, The Global Nutrition Cluster, CALP Network, and World Vision.

| Role | Organization |
|---|---|
| Head of Nutrition Department | Somalia Federal Ministry of Health (MoH) |
| Director | Federal Ministry of Labour and Social Affairs |
| SUN Movement Lead | Office of the Prime Minster |
| Somalia National Nutrition Cluster Lead | UNICEF |

| | |
|---|---|
| Somalia National Nutrition Cluster Deputy Coordinator | WFP |
| Director of Somalia Cash Working Group | Concern Worldwide |
| Director of Program Development and Quality | Save the Children International, Somalia |
| Head of Child Poverty Reduction | Save the Children International, Somalia |
| Global Goal Team Leader - Safety Nets and Resilient Families | Save the Children International (Centre) |
| Evidence for Impact Director | Save the Children US |
| Head of Cash and Voucher Assistance (CVA) | Save the Children International (Centre) |
| Nutrition Advisor | USAID/Bureau for Humanitarian Assistance (BHA) |
| Senior Nutrition Adviser (Concern Worldwide)<br><br>Nutrition Specialist (UNICEF) and co-lead of GNC | The Global Nutrition Cluster |
| ESAF Regional Representative | CALP Network |
| ESAF Deputy Regional Representative | CALP Network |
| Impact Evaluation Analyst | World Food Programme |
| Data and M&E Specialist \| Social and Behaviour Change | UNICEF |
| Sr Adv in Design, Evaluation and Research in Nutrition | World Vision |

### Intended outcomes of engagement with priority stakeholders

| Stakeholder name or group | Intended change outcome | Stakeholder quadrant (at start) | Indicator/s of success at end of study | Sources of evidence for the indicators |
|---|---|---|---|---|
| **Somalia MoH and NBS** | Work in partnership and Persuade | Develop interest | Focal points provide letters of support and engage meaningfully at each stage of the research<br>Support with obtaining ethical approval | • Email from MoH and NBS<br>• Letters of support from MoH and NBS<br>• Letter of IRB Approval<br>• MoH and NBS participation in the |

| | | | | |
|---|---|---|---|---|
| | | | Engagement with other MoH officials and sharing results of the research within the MoH<br>MoH uses evidence to inform the National Health Strategy Refresh | Research Kick-off in Nairobi<br>• MoH participation (in-person or virtual) in research dissemination event<br>• MoH consultants SC technical advisors during Strategy Refresh process |
| **World Food Programme (WFP)** | Work in partnership | Develop interest | WFP Somalia agrees to fund 3 additional months of implementation and partners with SC in the research study | Formal agreement (MOU) |
| **National Nutrition Cluster engagement** (MoH Nutrition Dept, WFP, UNICEF) | Persuade | Develop interest, capacity | Invitation to present research findings at one their Monthly Meetings<br>Cluster leads will disseminate summary of findings (research briefs) via email with partners<br>Uses research findings to inform the cluster-level decisions related to implementing various nutrition-specific and sensitive complementary activities in cash programming | • Email invitation to the Monthly Meeting<br>• Email with research brief attachments shared with Cluster partners |
| **Save the Children SNRF** | Work in partnership | Develop interest, Persuade | Invitation to present research findings at internally and externally | • Email invitation to the SNRF Quarterly Webinar<br>• Email with research brief attachments shared with SC senior leadership |
| **Somalia National Institute of Health (NIH)** | Monitor | Develop interest | Participation in the Second Annual National Health Conference in February 2024 | Email confirmation of abstract selection for and invitation to the conference |
| **SUN Movement** | Work in partnership, Persuade | Develop interest, | Establishment of formal agreement on advocacy of nutrition- | Memorandum of Agreement (MOU) |

| | | | | |
|---|---|---|---|---|
| | | strengthen partnership | sensitive programming and future research across Somalia<br><br>Support dissemination of our research findings<br><br>Advocacy/fundraising for future nutrition research | |
| Journal of Global Heatlh | Work in partnership | Develop interest | Publish journal collection with main findings and sub-study results | Email confirmation of interest in publishing Somalia wasting-focused journal collection |
| Head of CVA, Save the Children International | Work in partnership, Persuade | Develop interest | Fund a global, donor-facing cash learning event to share and discuss key findings and lessons from study to fundraise for future research. | In-person Learning Event |
| Nutrition Advisor, USAID/BHA | Monitor, Persuade | Develop interest | Share findings to generate further interest from donor to continue funding CVA + Nut in Somalia and use evidence from this study to advocate funding for other contexts (implementation and research). | |
| Global Nutrition Cluster (GNC) | Work in partnership, Persuade | Develop interest | The Global Nutrition Cluster will uptake the findings of the research, disseminate within its members and promote at national- and global-level learning and evidence-based approaches. | Webinars organized by GNC to present findings of the research |

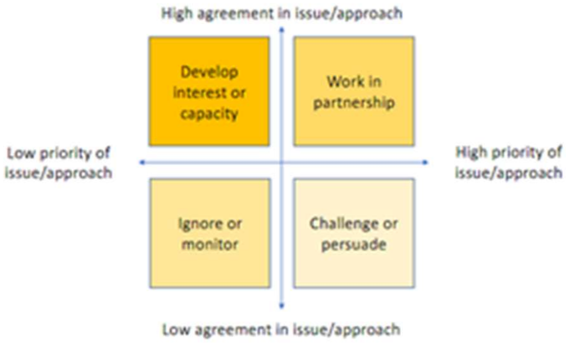

### Communications analysis of priority stakeholders

| Stakeholder name or group | Intended change outcome | Key messages | Preferred channels | Formats | Language |
|---|---|---|---|---|---|
| National MoH | Secure partnership and support for research | - The research aligns with the MoH's objectives<br>- We are interested in partnering with the MoH as our local research partner<br>- Interested in MoH supporting the research uptake within the Ministry | Emails<br><br>1:1 Meetings<br><br>Participation in various research activities (Kick-off, dissemination, etc) | Short presentations | English |
| National Health of Institute (NIH) | Secure support for dissemination | - The research is filling a critical gap in the global and national evidence around cash and nutrition-sensitive programming, particularly on relapse | Emails<br><br>1:1 Meetings | Short presentations | English |
| National Nutrition Cluster | Secure support for dissemination, inform national advocacy policy, and mobilization of further research | - The research is filling a critical gap in the global and national evidence around cash and nutrition-sensitive programming, particularly on relapse | Emails<br><br>1:1 Meetings<br><br>Monthly Cluster Meetings | Short presentations<br><br>Partner discussions | English |

| | | | | | |
|---|---|---|---|---|---|
| | | - Use the research to advocate and scale up evidence-based nutrition-sensitive programming across Somalia | | | |
| The Global Nutrition Cluster | Secure support for dissemination at the global level and influence guidance on designing cash-based programs for nutrition | - The research is filling a critical gap in the global evidence around cash and nutrition-sensitive programming<br>- Use the research to scale up evidence-based nutrition-sensitive programming globally and provide operational guidance on activities and duration of intervention | Emails<br><br>1:1 Meetings<br><br>Monthly Cluster Meetings | Short presentations<br><br>Partner discussions | English |
| Somalia Cash Working Group | Secure support for research, dissemination and adoption of findings into cash programming in Somalia | Use the research to scale up evidence-based nutrition-sensitive programming in Somalia and influence cash transfer values and frequency based on research | Emails<br><br>1:1 Meetings<br><br>Monthly WG Meetings | Short presentations<br><br>Partner discussions | English |
| SUN Movement | Secure support for dissemination across various sectors (Ministry of Ag, etc., Ministry of Social Affairs, etc.) | - The research is filling a critical gap in the global and national evidence around cash and nutrition-sensitive programming, particularly on relapse<br>- Use the research to advocate and scale up | Emails<br><br>1:1 Meetings | Short presentations<br><br>Partner discussions | English |

| | | |
|---|---|---|
| | | evidence-based nutrition-sensitive programming across Somalia |

### Engagement milestones

| Y | Q | Date/Location/Event | Engagement /communications milestone deliverable |
|---|---|---|---|
| **2023** | Q1 | *March: Formation of Research Committee, online* | *Ongoing weekly/bi-weekly meetings with research team (including government, JHU and SC broader team) to discuss research design, data collection implementation, challenges/solutions and research uptake/dissemination* |
| | Q2 | *April 27-May 1: Somalia R2HC Kick-off Workshop, Nairobi* | • *Participants gain a broad understanding of the research aims, objectives, and proposed study design.*<br>• *Discuss and agree on the timeline to finalize the study protocol including methodology, and tools.*<br>• *Introduce study stakeholders to each other, discuss roles and ways of working; amend project workplan.*<br>• *Identify and help find solutions to existing and future challenges.*<br>• *Discuss stakeholders' engagement and identify key activities for SES and research uptake plan.* |
| | Q3 | *August 3: Baseline Results Presentation and Discussion, online*<br><br>*August 9: Stakeholder Engagement and Research Uptake Planning, online* | *Share preliminary findings with Research Committee and Save the Children Global CVA and Nutrition teams*<br><br>*Prioritize, plan, and cost activities and products mapped during the Kick-off workshop. Assign roles and responsibilities across the Research Committee and update the SES.* |

| | | | |
|---|---|---|---|
| | Q4 | *November 29: Midline Results Workshop, online and in-person*<br><br>*December 5: WFP Impact Evaluation Forum, in-person, Rome* | *Share preliminary midline findings against final baseline findings with Research Committee, local research partners (MoH, NBS), and SC Global CVA and Nutrition teams*<br><br>*Present the research study methodology on the 'Cash-based Programming Design' Panel* |
| **2024** | Q1 | *March 13: J-PAL Training on Randomized Impact Evaluations for Humanitarian Professionals* | Present the RCT, namely challenges. |
| | Q2 | *April 28-30: Somalia R2HC Close-out Workshop, Nairobi*<br><br>*April 29: Conducting Impact Evaluations of Adaptive Social Protection in Fragile Settings: Design Characteristics and Methodologies* | *To discuss study results, explore key implications, conclusions, and recommendations. Plan dissemination events and products.*<br><br>*Share experience designing cash interventions and the Somalia RCT at the UNICEF Workshop.* |
| | Q3 | *July 23: Research Findings Presentation to Elrha – virtual* | *Present the final research findings and recommendations to donors (implementation and research)* |
| | Q4 | *Begin Research Dissemination (publications, conferences, workshops, webinars, etc.)* | *Several products developed targeting priority stakeholders (e.g., implementers, donors, researchers, and policymakers).*<br><br>*Written materials to be shared via *academic journal publications and research briefs; findings shared via SC and JHU social media platforms and internal bulletins; uploaded to CALP Library; shared at Global Nutrition Cluster member meeting; and other targeted dissemination such as donors).*<br><br>**Journal Supplement submission to JoGH:*<br>• *Paper 1: Main Study (Main results, cost-effectiveness and qualitative)*<br>• *Paper 2: Protocol paper*<br>• *Paper 3: Drivers of Wasting paper (Assessing the Drivers of Wasting* |

| | | | |
|---|---|---|---|
| | | | *among Children Under 5 and Their Mothers in the Bay and Hiraan Regions of Somalia)*<br>• *Paper 4: Measurement concordance paper (Analyzing concordance between MUAC, MUACZ, and WHZ in diagnosing acute malnutrition among children under 5 in Somalia)*<br>• *Paper 5: Relapse paper*<br>• *Paper 6: Commentary on the way forward*<br>**September to December**: abstract submissions to the International Congress of Nutrition (ICN); the Agriculture, Health, and Nutrition Academy Conference (AHN); and the Elrha Research Forum. |

**Table S14. Mobile Messages Received by BHA Participants**

| Somali Text | English Translation |
|---|---|
| *Maka warqabta Muhiimada ey u leedahay nafaqeynta wanaagsan dhalaanka iyo caruurta yaryar?* | Are you aware of the importance of good nutrition for your infants and young children? |
| *Waxannu Nuujina dhallaankeena si baraxtiran muddo lix bilood ah sababto ah Caanaha naaska wa cuntada ugu haboon dhallaanka waxeyna siiyan dhaman tamarta iyo nafaqooyinka u ilmaha u baahanyahay billaha ugu horeeya e noloshiisa.* | We breastfeed our babies exclusively for 6 months because breastmilk is the ideal food for infants and provides all the energy and nutrients that the infant needs for the first months of life. |
| *Waxeynu nuujina naaska ilaa caruurteena ey laba sana jirsadaan sababto ah labada sano ee ugu horeyso wa fursada caruurteenu ey ku goraan uguna diyaar garooban aduunka.* | We breastfeed until our children are two years old because the first two years is our children's opportunity to grow and be ready for the world. |
| *Waxaan nuujina naaska sababto ah caanaha naaska wa badbaado, nadiif ah o ka gooban unugyada difaaca jirka kawaaso ka hortago cuduro badan oo ku dhaca ilmaha yaryar.* | We breastfeed because breastmilk is safe, clean and contains antibodies that help protect against many common childhood illnesses. |
| *Xasuusnow inad ku dhaqanto nadaafada iyo fayadhowrka wanaagsan si ay kaga caawiso ka hortaga jirada o ad u hubiso caafimad qabka ilmahaaga.* | Remember to practice good hygiene and sanitation to help prevent illness and ensure your child stays healthy. |
| *La hadal adeeg bixiyaha caafimadka iyo shaqaalaha caafimadka e bulshada si aad wax badan uga ogaato quudinta haboon e dhalaanka iyo caruurta yaryar iyo sidi aad u siin karto ilmahaaaga bilowka ugu wanaagsan e nolosha.* | Talk to your healthcare provider or community health worker to learn more about IYCF and how you can provide your child with the best possible start in life. |
| *Si wadajir ah waxan siin karna caruurteena hadiyad caafimad o wanaagsan iyo mustaqbal ifaaya, ha ka habsaamin e bilow dhaqan galinta quudinta wanaagsan e dhalaanka iyo ilmaha yaryar.* | Together, we can give our children the gift of good health and a brighter future. Don't wait, start practicing good Infant and Young Child Feeding practices today. |
